# Fostering transparent medical image AI via an image-text foundation model grounded in medical literature

**DOI:** 10.1101/2023.06.07.23291119

**Authors:** Chanwoo Kim, Soham U. Gadgil, Alex J. DeGrave, Zhuo Ran Cai, Roxana Daneshjou, Su-In Lee

## Abstract

Building trustworthy and transparent image-based medical AI systems requires the ability to interrogate data and models at all stages of the development pipeline: from training models to post-deployment monitoring. Ideally, the data and associated AI systems could be described using terms already familiar to physicians, but this requires medical datasets densely annotated with semantically meaningful concepts. Here, we present a foundation model approach, named MONET (**M**edical c**ON**cept r**ET**riever), which learns how to connect medical images with text and generates dense concept annotations to enable tasks in AI transparency from model auditing to model interpretation. Dermatology provides a demanding use case for the versatility of MONET, due to the heterogeneity in diseases, skin tones, and imaging modalities. We trained MONET on the basis of 105,550 dermatological images paired with natural language descriptions from a large collection of medical literature. MONET can accurately annotate concepts across dermatology images as verified by board-certified dermatologists, outperforming supervised models built on previously concept-annotated dermatology datasets. We demonstrate how MONET enables AI transparency across the entire AI development pipeline from dataset auditing to model auditing to building inherently interpretable models.

## Introduction

Ensuring the transparency and robustness of medical AI systems involves assessing data and models at every stage, from model training to post-deployment monitoring. However, the tools and methods needed to promote AI transparency and to de-mystify “black-box” models often require medical datasets with dense annotations of human-understandable concepts. For example, for building a melanoma classifier, it would be medically meaningful to understand the data and model using concepts such as “darker pigmentation”, “atypical pigment networks”, and “multiple colors”. Unfortunately, obtaining such labels requires a significant amount of time from domain experts, and consequently, most medical datasets limit annotations to little more than diagnoses. In contrast, *rich annotation* with the extensive and highly descriptive clinical concepts developed by the medical community could enable numerous benefits. Such rich annotations could promote understanding of key biases in datasets, empower detection of undesirable behavior in medical AI devices, and foster the development of AI devices that better align with physicians’ expectations. However, few medical image datasets include such extensive annotations, and the time expended in existing efforts [1] argues that obtaining this data via large-scale efforts by human experts is infeasible.

Here, we instead leverage the collective knowledge of the medical community, as encapsulated in publicly available medical literature and medical textbooks, to teach an AI model, MONET (**M**edical c**ON**cept r**ET**riever), to richly annotate medical images with semantically meaningful and medically relevant concepts (Fig. 1A-B). We focus on the application of dermatology to showcase its versatility since dermatology has heterogeneity in disease appearance across diverse skin tones and has no standardized imaging practices, leading to significant heterogeneity in imaging conditions (*e.g.,* lighting, blurriness). In this setting, examples of clinical concepts include lesion color (*e.g.,* brown) and morphology (*e.g.,* nodule). MONET’s automatic concept generation capability empowers us to perform meaningful trustworthiness analysis across all stages of the medical AI pipeline, as demonstrated by three use cases (Fig.1C-E).

**Fig. 1.**
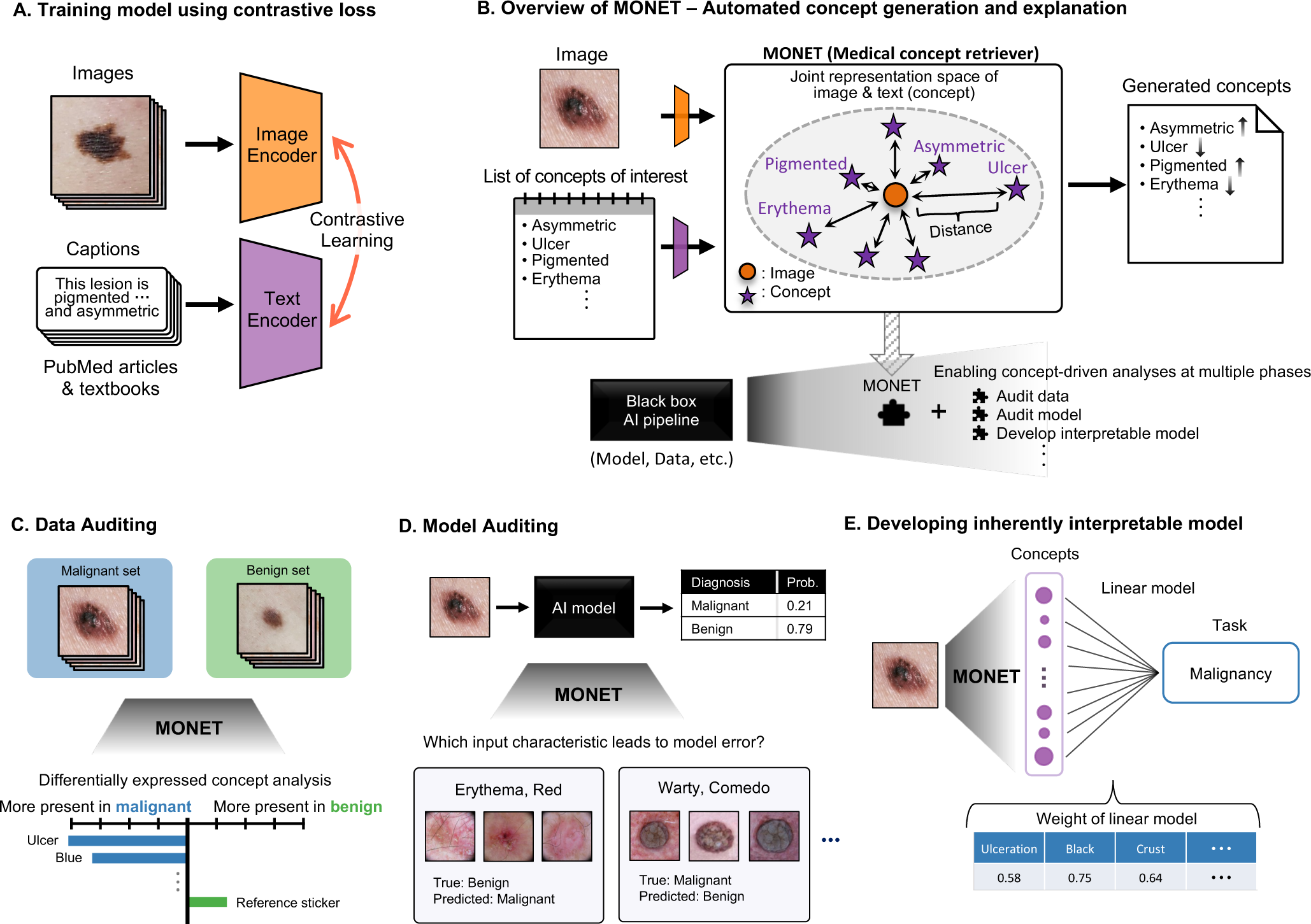
Overview of MONET framework and its usage examples. **(A)** *Training procedure.* MONET is trained using contrastive learning on an extensive set of dermatology image and text pairs collected from PubMed articles and medical textbooks. During the training process, the paired image and text are forced to be close in the joint representation space, while those from different pairs are forced to be far apart. **(B)** *Automatic concept generation.* MONET can map medical concepts and images onto a joint representation space, allowing it to determine the degree to which a concept is present in an image for any given concept by measuring the distance between the image and concept text prompts in the representation space. Its concept generation capability enables various concept-driven analyses at multiple stages of the medical AI pipeline. **(C)** *Concept-level data auditing.* MONET’s automatic concept generation capability makes it possible to explain the distinguishing features between two sets of data in the language of human-interpretable concepts. This approach facilitates the auditing of large-scale datasets with ease. **(D)** *Concept-level model auditing.* MONET can be used to identify which input characteristic leads to the errors of medical AI. **(E)** *Developing inherently interpretable models.* MONET can be used to develop inherently interpretable medical AI models that operate on human-interpretable concepts aligning with physicians’ expectations. These models allow physicians to easily decipher the factors influencing the models’ decisions, ensuring high transparency.

Dataset auditing can identify biases in the data before using it for any clinically relevant task, thereby improving the quality of the data, providing an opportunity for preliminary bias mitigation, and improving overall trustworthiness in the data. Prior work in dataset auditing has identified how particular concepts are associated, either appropriately or inappropriately, with data labels [2–4]. In medicine, data auditing has identified spurious correlations in AI training data [5, 6]. For example, the overrepresentation of chest drain in the x-rays of patients with pneumothorax led to AI algorithms that relied on their presence. However, chest tubes are a treatment used after a physician had diagnosed pneumothorax and not a causal feature [5]. Because data is not static, dataset auditing also allows the detection of dataset shifts or drifts by identifying the changes in the representation of a concept in the data [7–9]. MONET enables us to examine datasets on the basis of a rich set of automatically retrieved concepts (Fig.1C).

Model auditing involves demystifying the “black-box” of AI models – understanding the factors involved in AI decision-making [10–13]. AI models that fail during real-world deployment can lead to worse outcomes for patients. AI models have been shown to make systematic errors on a subset of data with shared features, resulting in uneven performance across the data [2–6, 13, 14]. To prevent this and make appropriate adjustments, models should be audited to understand their failure modes prior to deployment. Model auditing is not only important immediately after model development, but is a continual process, especially since models may undergo updating over time [7–9]. MONET powers model auditing: the dense concept annotations generated by MONET can be used to understand which input characteristic leads to model errors (Fig.1D).

While most existing AI models are black boxes, newer methods in the field of explainable AI have attempted to create inherently interpretable models that use concept-level features as input [15, 16]. The automatic generation of a rich set of semantically meaningful concepts allows us to leverage and enhance these recent models (Fig.1E).

Because of the need to develop and test the aforementioned methods and tools, there have been prior attempts to create densely annotated datasets in medicine. Such datasets include SkinCon [1], PH2 [17], derm7pt [18], and Osteoarthritis Institute Knee X-ray dataset (OAI) [19]. However, because these medical datasets are annotated by humans and usually require domain expertise, the number of densely annotated datasets, the number of images in each dataset, and the number of “concepts” that can be labeled in each image are limited. MONET overcomes these challenges by generating medical concepts automatically. Our framework is built upon *contrastive learning*, a recent AI breakthrough that enables the direct utilization of natural language descriptions on images [20]. Since this approach does not require manual labeling, it can unlock the potential of vast numbers of image and text pairs, allowing for the harnessing of data of much larger scale than was possible with supervised learning.

To train MONET, we collect an extensive set of dermatology image and text pairs (*n* = 105, 550) from PubMed articles and medical textbooks. We map an image (or text) into a lower-dimensional vector or a *representation* through a neural network, namely the *encoder* (Fig.1A), creating a *representation space*. During the training process, an image and text from the same pair are forced into close proximity in the representation space, while those from different pairs are forced to be farther apart (Methods). Once trained, MONET’s *zero-shot capability* (*i.e.,* the ability to generate a medical concept without a separate learning procedure) generates concepts (Fig. 1B and Methods). When a user provides an image and a list of concepts to generate, MONET determines the presence of each concept in the image by calculating the distances between the image and concept text prompts in the joint representation space, where images and texts are jointly mapped.

MONET’s automatic concept generation capability enables a whole range of capabilities in medical AI that were previously infeasible in practice. We showcase MONET’s versatility by demonstrating its use in auditing data, auditing models, and creating inherently interpretable models. MONET enables a sophisticated multi-point analysis and can be used to probe any part of the medical processing workflow. For data auditing, we apply MONET to the International Skin Imaging Collaboration (ISIC) dataset [21–26], the most widely used data in dermatology AI [27], to confirm known trends and discover new ones. We also use MONET to identify which input characteristics lead to errors in medical AI models. Finally, we integrate MONET with the concept bottleneck model (CBM) [15], a well-known approach for building inherently interpretable models, and show MONET+CBM’s advantages over supervised models in terms of *both* performance and interpretability. All of these tasks are central to the development and deployment of trustworthy and transparent AI models in medicine.

## Results

### Automatic concept generation

We first assess MONET’s concept generation capability before demonstrating how this capability can improve the transparency and interpretability of medical AI pipelines. The fundamental mechanism in MONET’s concept generation is the mapping of medical concepts and images onto a joint representation space. This allows the generation of a *concept presence score, i.e.*, the degree to which a concept is present in an image, by measuring the distance between the image and concept text prompts in the joint representation space (Methods). We evaluate MONET’s concept generation ability by identifying those images with the highest concept presence scores using both *clinical* and *dermoscopic* images, the two widely used dermatological image types. Dermoscopic images are captured using digital photography with a specialized dermatological instrument called a dermoscope that magnifies skin lesions to capture fine details, while clinical images are often taken at least 6 cm away with a digital camera. For our evaluation, we employ clinical images (*n* = 4, 960) from the Fitzpatrick17k and Diverse Dermatology Images (DDI) datasets and dermoscopic images (*n* = 71, 242) from the ISIC dataset (Methods).

Fig. 2 and Supplementary Fig. 1-2 display clinical and dermoscopic images with high concept presence scores for each concept. These represent examples of widely used medical concepts in dermatology. Dermatologists use a standardized terminology to describe the morphology, color, configuration and distribution of skin lesions. MONET excels at recognizing these medical concepts in clinical and dermoscopic images. For example, “erythema” is a term used by dermatologists to describe a red or violaceous color, which usually occurs in the presence of inflammation. It can be found in various skin diseases, such as atopic dermatitis, psoriasis, and rosacea. Two board-certified dermatologists confirmed that the images with large presence scores for erythema exhibit skin redness in both clinical and dermoscopic images (Fig. 2). Similarly, images with the concept “blue” show dark blue lesions with pigmentation in the dermis, resulting from the Tyndall effect. Moreover, MONET was able to retrieve images with primary morphological features such as bullae (large, tense fluid-filled blisters) and pustules (small, pus-filled blisters), as well as secondary morphological features including ulcers (open sores) and xerosis (dry, scaly skin).

**Fig. 2.**
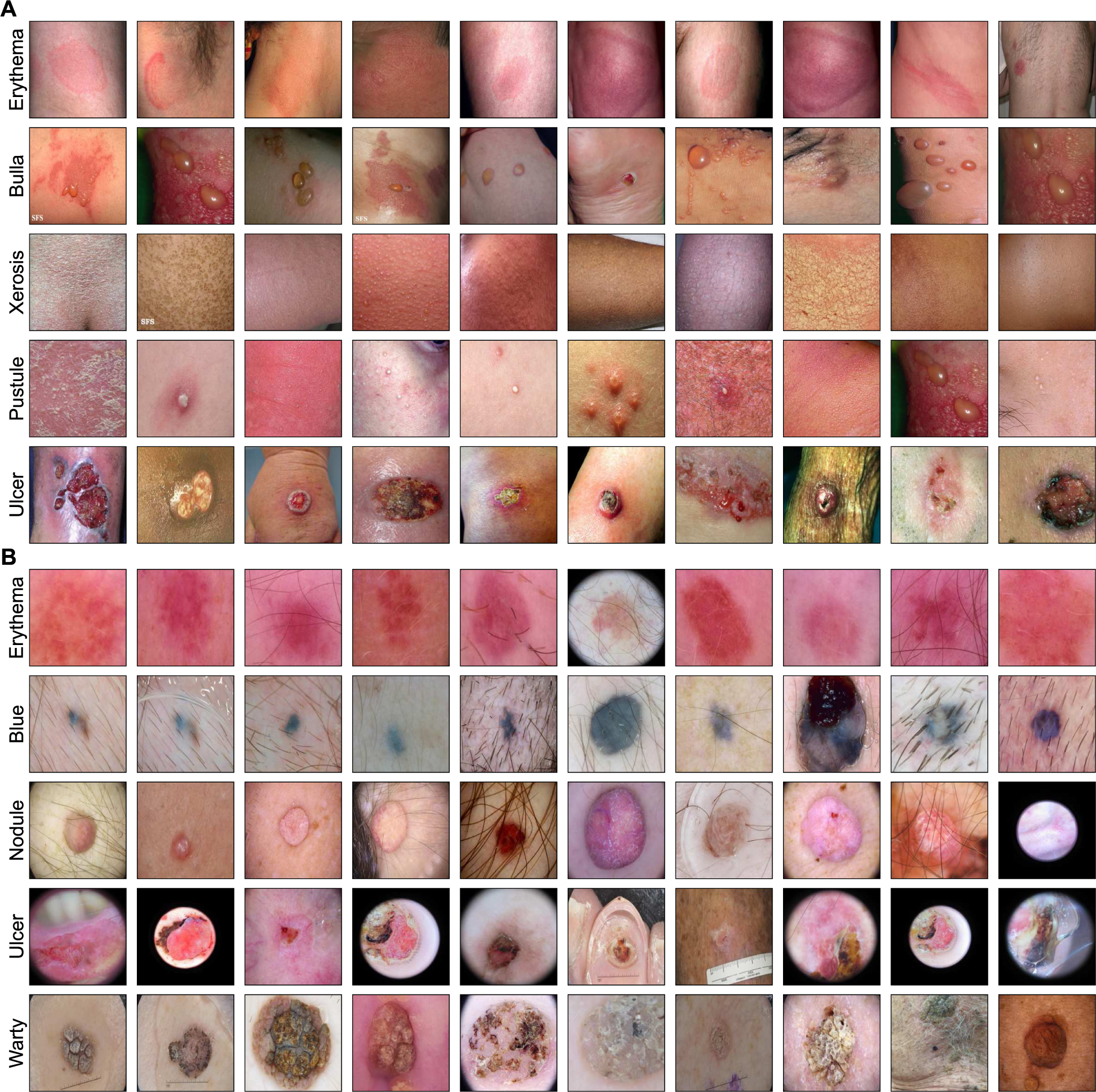
Images with high concept presence scores calculated using MONET. The concept presence score represents the degree to which a concept is present in an image. Each row displays the top 10 images for each concept. **(A)** Clinical images from the Fitzpatrick17k and DDI datasets. We exclude images inappropriate for public display due to the inclusion of sensitive body parts; for completeness, we denote the filenames of these files in Supplementary Table 1 **(B)** Dermoscopy images from the ISIC dataset.

We assess the performance of MONET’s concept generation using ground truth concept labels in SkinCon (Table 1). Of the 48 concepts in the dataset, we exclude any with less than 30 positive examples, leaving 21 concepts for our analysis. We use 1,645 images from Fitzpatrick17k and DDI datasets with ground truth SkinCon concept labels. We compare MONET’s performance to a supervised learning approach, training a ResNet-50 model using ground-truth concept labels from SkinCon [28], and to a pre-existing contrastive image-text model that was not specifically trained on dermatology images but on 400 million available image-text pairs on the web - the CLIP (Contrastive Language-Image Pretraining) model by OpenAI [20] (Methods). We find that MONET outperforms the ResNet-50 model and the CLIP model in terms of concept generation. Specifically, we compare the mean of the area under the receiver operating characteristic curve (AUROC) across concepts with ground truth labels and count how many concepts achieved an AUROC higher than 0.7. MONET achieves a mean AUROC of 0.766; in contrast, CLIP achieves a mean AUROC of 0.692. The ResNet-50 model, trained to predict concept labels, achieves a mean AUROC of 0.692. Of the 21 concepts analyzed, MONET remarkably displays 19 concepts with an AUROC over 0.7, compared to 9 for CLIP and 11 for the fully supervised model. Additionally, we conduct the same comparative analysis using disease labels, which can be viewed as the most fine-grained concept labels; we map the disease labels instead of SkinCon concepts to the image-text joint representation space. Our findings indicate that MONET’s performance is still comparable to that of supervised models in this case (Supplementary Table 2). This observation is consistent with a previous study, which found that a contrastive learning model trained on radiology images demonstrates comparable performance in predicting pathologies in chest X-rays to that of supervised learning models [29].

**Table 1.** Performance of MONET’s concept generation as compared to the baselines. We use concept labels in the SkinCon dataset as ground truth. Of the 48 concepts in the dataset, we exclude any with less than 30positive examples, leaving 21 concepts for our analysis. The baselines are CLIP, an image-text model not specifically trained on dermatology images, and the ResNet-50 model trained on ground truth labels in a fully supervised manner. The numbers in parentheses represent the count of concepts for which the method achieves an AUROC over 0.7 over the total number of concepts examined.

| Method | Mean AUROC |
| --- | --- |
| MONET | 0.766 (19/21) |
| CLIP | 0.692 (9/21) |
| ResNet-50 (Fully supervised) | 0.692 (11/21) |

We also evaluate the performance of MONET’s concept generation across diverse skin tones (Methods). A recent study revealed that state-of-the-art dermatology AI models exhibit uneven performance across skin tones, particularly underperforming on dark skin tones, potentially due to the insufficient representation of diverse skin tones in training data [30]. One advantage of contrastive learning, the technique used to train MONET, is its ability to easily harness heterogeneous data from diverse sources for training. This can help reduce performance disparities across demographics compared to training on a single data source. To determine whether MONET is free from this issue, we compared its performance per skin tone using the Fitzpatrick skin type labels included in the Fitzpatrick17k and DDI datasets. MONET demonstrated even performance across skin tones (Supplementary Table 3).

Finally, we also explore MONET’s capability to recognize non-clinical concepts, such as artifacts that are irrelevant to the diagnosis. Many studies have shown that medical AI systems use such non-clinical concepts to make predictions, particularly when a spurious correlation exists between the artifacts and prediction labels [6, 31]. In dermatology AI, it has been shown that artifacts, such as clinical marking or size reference stickers, can have a detrimental effect onthe model’s generalizability [32–34]. The ability of MONET to identify such artifacts, in addition to clinical concepts, will facilitate more fine-grained auditing and debugging of medical AI pipelines. Supplementary Fig. 3 shows images from the ISIC dataset that MONET identified as containing non-clinical concepts. Supplementary Fig. 3A shows images with purple pen ink marking that MONET automatically identified; in dermatology, lesions that are biopsied are often routinely marked with purple ink markers. Supplementary Fig. 3B shows orange stickers that MONET identified; they serve as a lesion marker. In ths ISIC dataset, these orange stickers predominantly show up in the pediatric cases, which are largely benign. As these artifacts predominantly appear in certain types of images, they may inadvertently cause AI algorithms to associate purple ink markings with malignancy and orange stickers with benign lesions [34]. Also, MONET automatically identifies images with body location features (such as nails and hair) (Supplementary Fig. 3C, D). A recent study has shown that anatomic locations may play a critical role in the performance of dermatology AI algorithms; however, most datasets lack these annotations [35]. Further,MONET identifies images with dermoscopic borders, which appear on a subset of dermoscopic images depending on the image processing process (Supplementary Fig. 3E).

In the following sections, we showcase how MONET can be used to improve the transparency and trustworthiness of dermatology AI in real-world scenarios.

### Data auditing

Ensuring that training data aligns with users’ expectations is a crucial first step toward developing AI models since many unreasonable model behaviors stem from unidentified pitfalls in the training data [6, 32, 34]. For example, in dermatology, when preparing a dataset for training an AI model to diagnose malignancy, the differentiating features in the data between classes (*i.e.,* malignant and benign images) should not contain any biases or spurious correlations, such as the pen markings used to identify biopsied lesions [34]. Upon identifying any irregularities, adjustments can be made, such as improving the data collection and processing [36, 37] or applying optimization techniques to improve generalizability [31, 38].

However, examining large-scale datasets for irregularities is challenging and labor-intensive. One approach is to manually label features of interest and create a contingency table between each feature and the target label to check for spurious correlations; however, this is subjective and not easily scalable [5]. Another approach is to train a generative model, such as CycleGAN [39], to learn the distribution of data for each class [6]; the trained generative model can modify an image from one class to resemble an image from another class. By observing these changes, a data examiner can identify the distinguishing features of each diagnostic group. However, generative models are difficult to train and necessitate manual inspection of the transformed images.

To address the issue, we can employ MONET to automate the data examination process. MONET’s automatic concept generation capability can explain the distinguishing features between any two arbitrary sets of images in the language of human-interpretable concepts, which we refer to as *concept differential analysis* (see Methods). Supplementary Fig. 4 shows benchmark analysis results.

As a practical use case, we employ MONET to analyze the ISIC dataset, the largest dermoscopic image dataset, which consists of over 70, 000 publicly available images that are commonly used to train dermatology AI models [21–27]. We divide the images into a malignant (*n* = 10, 091) and a benign set (*n* = 61, 151), assuming malignancy as the prediction target, and examine which concepts were more present in which set (Fig. 3A). We test for 48 concepts listed in SkinCon along with eight artifacts, including red coloration, pinkish coloration, and purple ink markings, nails, hair, orange sticker, gel, and dermoscopic border.

**Fig. 3.**
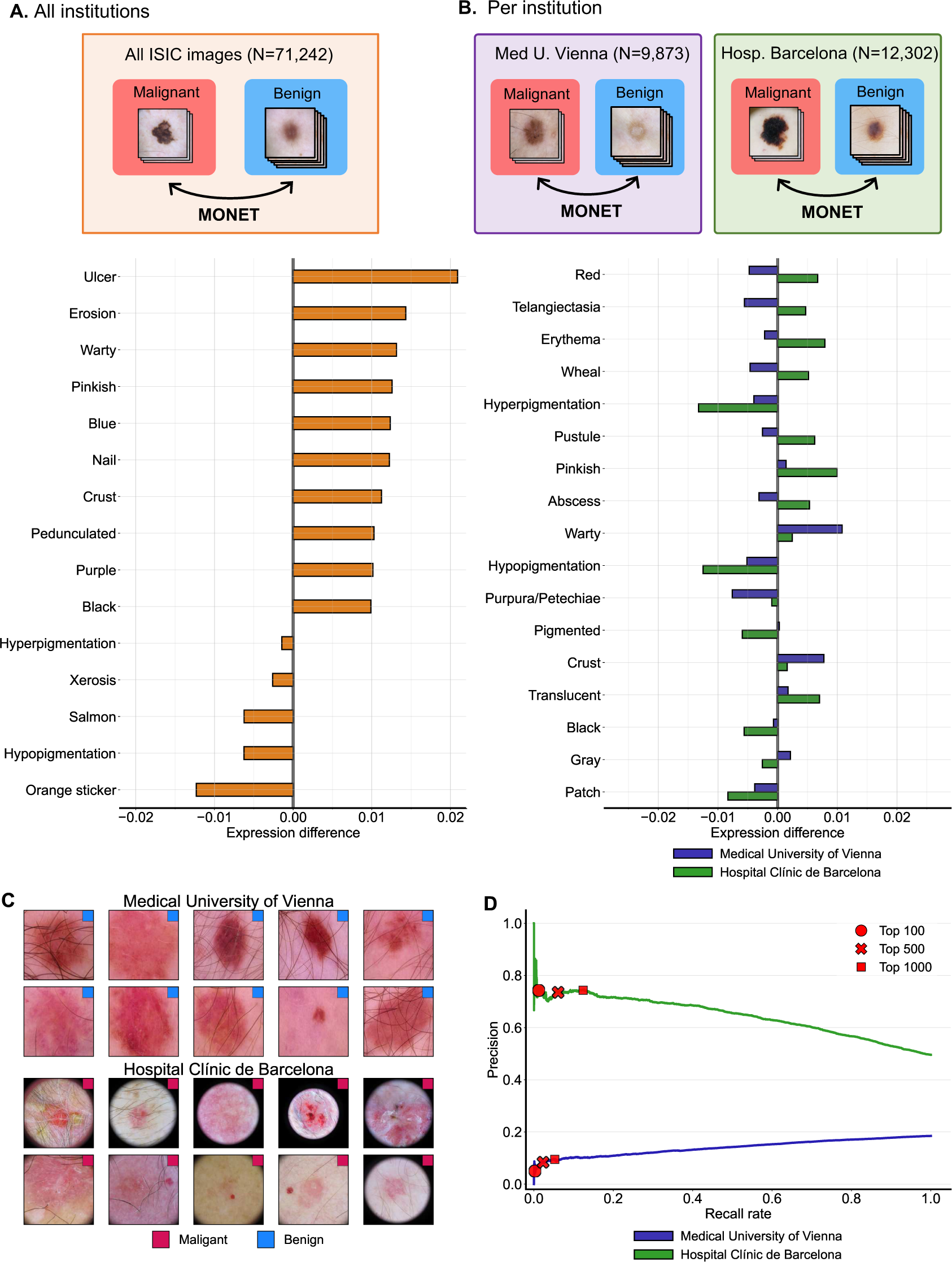
Concept-level data auditing. **(A)** We perform concept differential analysis between malignant images and benign images in the ISIC dataset. We show the top 10 concepts with positive values and the top 5 concepts with negative values. A positive value means the concept was more present in the malignant images than in the benign images, and vice versa. **(B)** We perform concept differential analysis between malignant and benign images per data source in the ISIC dataset to identify data-source-specific trends. The purple bar represents the output from the Medical University of Vienna, and the green bar represents the output from the Hospital Clinic de Barcelona. We show the top 15 concepts based on their absolute differences between the two cohorts. **(C)** Examples of red images in each cohort. We display 10 randomly selected images from the top 100 images in each cohort that had high concept expression scores for redness. **(D)** Precision-recall curve for images in each cohort. The images in each cohort are sorted based on their concept presence scores for redness and then compared to their malignancy labels. Precision is defined as the proportion of malignant images above a certain threshold out of all images above that threshold, while recall rate is defined as the proportion of malignant images above the threshold out of all malignant images. The top 500 and top 1000 red images from Barcelona Hospital still contain more malignant than benign samples.

The top 5 concepts in the malignant images are ulcer, erosion, warty, pinkish coloration and blue coloration; in contrast, the top 5 concepts in the benign images are orange sticker, hypopigmentation, the color salmon, xerosis, and hyperpigmentation. These concepts represent key features that a prediction model, trained on the ISIC dataset, may potentially use to differentiate benign from malignant lesions. They encompass both clinically pertinent features such as ulcer, crust, erosion, warty, and black coloration, as well as irrelevant confounders such as orange sticker and nail. Skin ulcerations and erosions are commonly linked to malignant skin tumors such as melanoma, basal cell carcinoma and squamous cell carcinoma, making their association with malignancy logical. On the other hand, prediction models learning from confounding concepts may lead to biases and detrimental consequences. For example, dermatologists use orange stickers as a lesion marker, and with the ISIC dataset, this was predominantly used in the pediatric population which had mostly benign lesions. The bias in the data could lead a model to erroneously associate orange stickers with a low likelihood of malignancy.

Furthermore, we can use this approach to assess distinctive trends specific to different data sources. In medicine, data sharing across institutions is limited due to the sensitive nature of medical data and regulatory constraints. In many cases, a medical AI is developed within a few institutions and then distributed to other institutions for deployment. For this reason, it is important to understand and monitor the shifts in the concept representations between data and identify factors that can potentially compromise the transferability of medical AI. By doing so, necessary adjustments can be made preemptively. The ISIC dataset is a collection of images from multiple hospitals and research institutions, which serves as an ideal resource for simulating situations where the development and deployment sites differ. In this analysis, we focus on two cohorts released in the ISIC Challenge 2019—the Medical University of Vienna (Med U. Vienna, malignant: *n* = 1, 824 / benign: *n* = 8, 049) and the Hospital Cĺınic de Barcelona (Hospital Barcelona, malignant: *n* = 6, 097 / benign: *n* = 6, 205)—since they represent the two largest cohorts in the entire ISIC dataset when stratified by release year and data source. We perform concept differential analysis between malignant and benign images, as noted above, for each cohort separately. We then compare the obtained concept differential expression scores between the two cohorts (Fig. 3B).

When we sort the test concepts in the order of absolute differences, the top-listed concept was a “red” hue. Redness is positively correlated with malignancy for the images from Hospital Barcelona but negatively correlated with malignancy for the images from Med U. Vienna. This means that redness has the potential to compromise the transferability of medical AI between two institutions. This trend is clearly visible in the sampled images from each cohort, as well. Fig. 3C displays images sampled from the top 100 images with high concept expression scores for the red coloration for each cohort, along with their diagnostic labels. The images that have more redness collected from Med U. Vienna are often benign, while those collected from Hospital Barcelona are often malignant. The top 500 and top 1,000 red images in the Med U. Vienna contain more benign than malignant ones, while the top 500 and top 1,000 red images from Hospital Barcelona still contain more malignant than benign samples. (Fig. 3D).

In sum, we demonstrate how MONET can assist with auditing large-scale datasets. Since concept differential analysis is conducted simply by describing a concept in a natural language, the approach fosters the scalable discovery of trends within the data. Using the insights gained through this process, AI model developers can enhance data collection, processing, and optimization techniques, ultimately yielding more reliable and trustworthy medical AI models.

### Model auditing

Various techniques for auditing AI models have been developed to understand the factors involved in AI decision-making. One classical approach is the use of *saliency maps*, which highlight regions in an input image that significantly contribute to the model’s prediction [40–42]. The saliency maps of each image help to identify which pixel-level features lead to a correct or incorrect prediction. However, the highlighted pixels are often not easily translated into semantically meaningful concepts understandable to a human [43].

To address this issue, we can use MONET to audit AI models through the lens of medical concepts. We developed a method “model auditing with MONET” (MA-MONET) that uses MONET to automatically detect semantically meaningful medical concepts that lead to model errors (Methods). MA-MONET starts by sorting images from a test set into groups based on their visual similarity. It then labels the clusters that perform below the overall accuracy as low-performing. For each low-performing cluster, MONET identifies medical concepts. Each low-performing cluster is then compared to a high-performing counterpart containing similar images, with concepts separately identified in the high-performing cluster. If two visually similar clusters (one high-performing, the other low-performing) differ in terms of a few concepts, these differing concept terms can be hypothesized as leading to high error rates. Finally, we produce a ranked list of medical concepts identified by MONET that differentiate the two clusters.

To validate our model auditing, we first perform a benchmarking analysis, using a situation where the ground truth (*i.e.,* the concepts leading to model error) is already known (Fig. 4A and Methods). We create a training dataset with spurious correlations from the Fitzpatrick17k and DDI datasets: 500 malignant images that feature aparticular SkinCon concept, while the 500 benign images do not. After training a CNN model to predict malignancy on this confounded dataset, we test it on a dataset where the correlation is reversed (Methods); in the test set, 500 sampled benign images have the SkinCon concept, while 500 sampled malignant images do not. We cluster images in the test set into 40 clusters, and about 20 of these clusters underperform, meaning their accuracy falls below the average accuracy. For these low-performing image clusters, we apply the MONET-based error explanation method to obtain a ranked list of medical concepts that would explain the model error. Finally, we observe if the concept of spurious correlation we know is recovered.

**Fig. 4.**
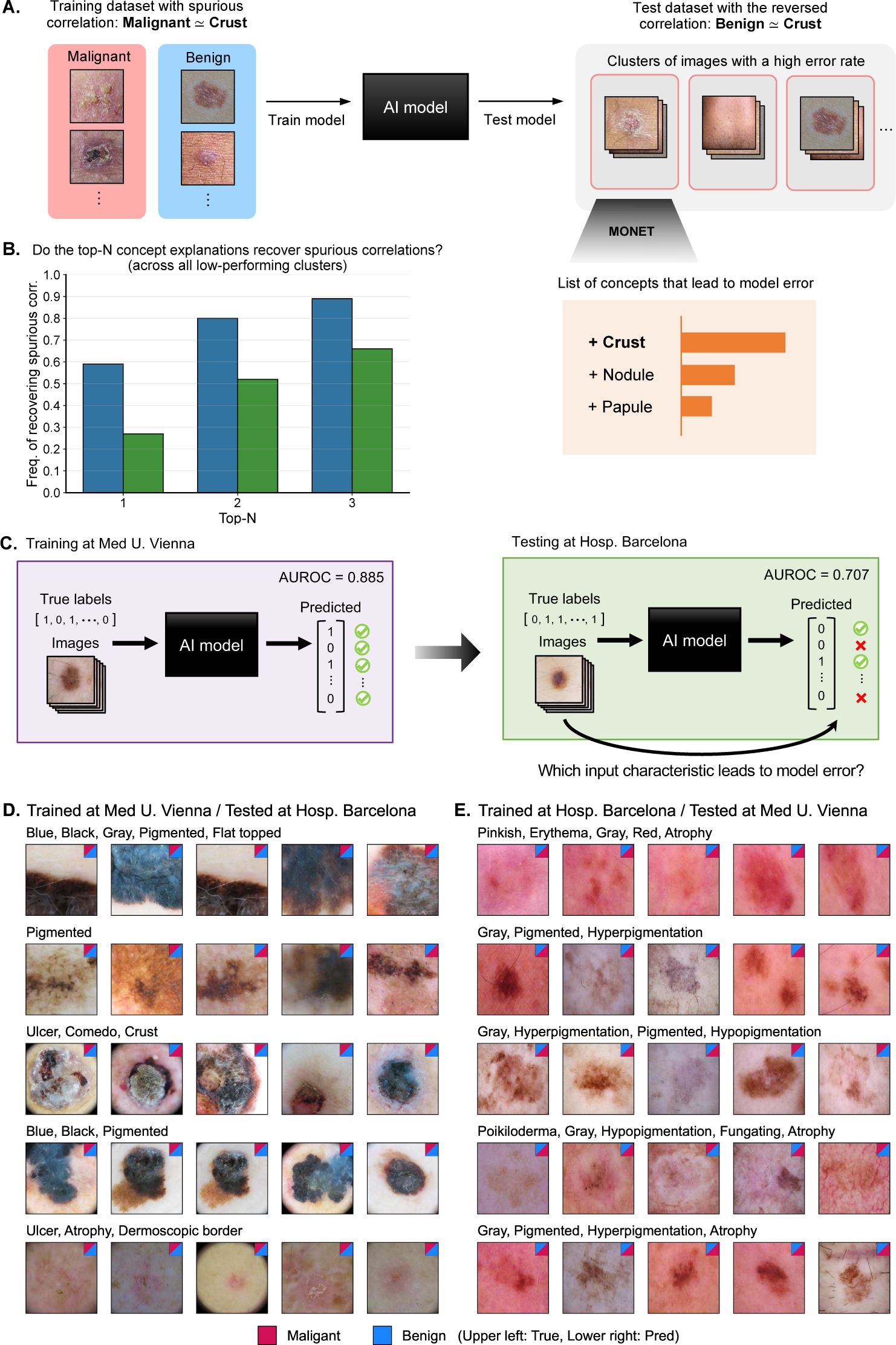
Concept-level model auditing. **(A)** We perform a benchmark analysis to see how well “model auditing with MONET” (MA-MONET) can identify the semantically meaningful concepts that lead to model error. To this end, we generate settings where we know the ground truth (*i.e.,* concepts that lead to model errors); we create a training and test dataset with spurious correlation. We use MA-MONET to identify which concepts lead to model error for an AI model trained on this confounded dataset. MA-MONET returns a ranked list of concepts that explain model errors. **(B)** The frequency of the known spurious correlation being recovered by MA-MONET is shown. **(D)**-**(E)** Each row displays one of the top 5 clusters, sorted by high error rates. For each cluster, we show the misclassified images and the corresponding concepts associated with errors. We represent the true and predicted labels for each image by the color of the upper left and lower right triangles in the small box, respectively. The numbers at the top right compare the number of malignant and benign samples for the true and predicted labels. The 5 misclassified images shown for each are selected based on the average concept presence of the identified concepts.

We conduct the analysis using 5 concepts that remain after filtering out concepts with fewer than 30 samples in each category required for creating the confounded training and test sets (*i.e.,* malignant–with concept, malignant–without concept, benign–with concept, and benign–without concept) : “crust”, “hyperpigmentation”, “plaque”, “erythema”, and “papule”. For each of the 5 concepts, we repeat this analysis 20 times with different random seeds changing the training and test sets. Consequently, we test 100 settings in total. Across these settings, the mean AUROC of the trained model is 0.779 for validation sets, but decreases to a mean of 0.458 for test sets.

We measure the frequency of the known spurious correlation being recovered by MA-MONET (Fig. 4B), checking if the top-N concept lists of any low-performing clusters include the known spurious correlation concept. We compare this outcome with that of the out-of-the-box CLIP model, which was not specifically trained on dermatology data [20]. The low-performing clusters being analyzed in each setting are the same for both methods, but the enumeration of concepts associated with errors is done using CLIP instead of MONET. The results for the top 1, 2, and 3 rankings are markedly higher with MONET, at 0.590, 0.800, and 0.890, respectively, compared to those obtained with CLIP, which are 0.270, 0.520, and 0.660, respectively.

To showcase its use in real-world scenarios, we consider a widely occurring situation where a model is trained at one institution and deployed at another [44, 45] (Fig. 4C). For training and testing, we use the same datasets we used in the data auditing section, specifically the two largest cohorts in the ISIC dataset: the Hospital Cĺınic de Barcelona (*n* = 12, 302) and the Medical University of Vienna (*n* = 9, 873). We train CNN models on the data from one institution using a standard training regimen and test them on the data from the other institution, and vice versa.

For the AI model trained on Med U. Vienna, it showed an AUROC of 0.885 in the internal validation, but the value dropped to 0.707 during the external validation. This decline in performance would prompt AI model developers to question which input characteristics led to model errors. To elucidate this, we use MA-MONET to pinpoint which concepts are associated with model errors. For each cluster with high error rates, the misclassified images and the terms associated with errors are shown in Fig. 4D (displaying the top 5 clusters sorted in the order of high error rates) and Supplementary Fig. 5 (displaying the top 15 clusters sorted in the order of high error rates). For example, the cluster with the highest error rate, displayed in the first row of Fig. 4D and Supplementary Fig. 5A, is characterized by the concepts “blue”, “black”, “gray”, “pigmented”, and “flat-topped”. Remarkably, we notice several clusters where high error rates are explained by concepts related to “red”. For instance, the cluster shown in Supplementary Fig. 5F are characterized by “erythema”, “salmon”, “sclerosis”, “scar”, and “translucent”. Interestingly, we also find that the malignant images are predominantly misclassified as benign. Out of the 74 malignant images in the cluster, 55 images are misclassified to be benign. This observation aligns with the trends we noted in the data auditing experiment, where red images from Med U. Vienna (*i.e.,* training dataset) were benign, while the red images in Hosp. Barcelona (*i.e.,* test dataset) were malignant.

Conversely, we also train an AI model on Hosp. Barcelona data and tested it on Med U. of Vienna data. Inthis case, the AUROC of 0.844 in internal validation drops to 0.741 during external validation. For each cluster with high error rates, the misclassified images and the identified terms associated with errors are shown in Fig. 4E and Supplementary Fig 6. The cluster with the highest error rate, shown in the first row of Fig. 4E, is characterized by the concepts “pinkish”, “erythema”, “gray”, “red”, “atrophy”. Out of the 197 benign images in the cluster, 103 images are misclassified as malignant. This observation also aligns with the trends we noted in the data auditing experiment.

### Inherently interpretable model building

In medicine, inherently interpretable models are of particular interest since they allow physicians to easily decipher the factors influencing a model’s decision. Rather than training a complex black-box model that requires post-hoc explanation, an inherently interpretable model offers greater transparency and control in model behavior. *Concept bottleneck models* (CBMs) are a well-known type of inherently interpretable model [15]. CBMs make predictions in a two-step manner: first, they predict concepts from the input using a complex model such as a CNN (*i.e.,* input → concept); then, they use these predicted concepts to predict the target output via a linear model (*i.e.,* concept → output). As each node in the bottleneck layer represents a human-interpretable concept, CBMs offer greater explainability. Further, CBMs facilitate the incorporation of domain knowledge into models by imposing constraints on the concepts used, thereby improving the ability to control model behavior.

However, CBMs have a significant limitation: they require concept annotations in the training data. To achieve high performance with CBMs, it is essential to train them on a sufficient number of samples and ensure they operate with an adequate number of concept labels that are relevant to the prediction task. This constraint has hindered the practical application of CBMs. We address this issue by using MONET’s automatic concept generation to eliminate the need for manually annotated concept labels in the original training procedure of the CBMs.

We explore the application of MONET and CBMs for melanoma and malignancy prediction tasks, the most prevalent prediction tasks in dermatology AI. MONET+CBM approach predicts the target (*i.e.,* melanoma or malignancy) by combining automatically generated concepts by MONET (*i.e.,* the concept presence score) via a linear model (Fig. 5A and Methods). This gives CBM access to many concepts and many samples compared to a manual labeling approach. The following comparison makes use of 4,960 clinical images sourced from the Fitzpatrick17k and DDI datasets. For melanoma prediction, we further filter images to ensure that data mirrors a well-defined clinical task, resulting in 775 images (Methods). For each setting, we repeated evaluations with 20 different train-test splits.

**Fig. 5.**
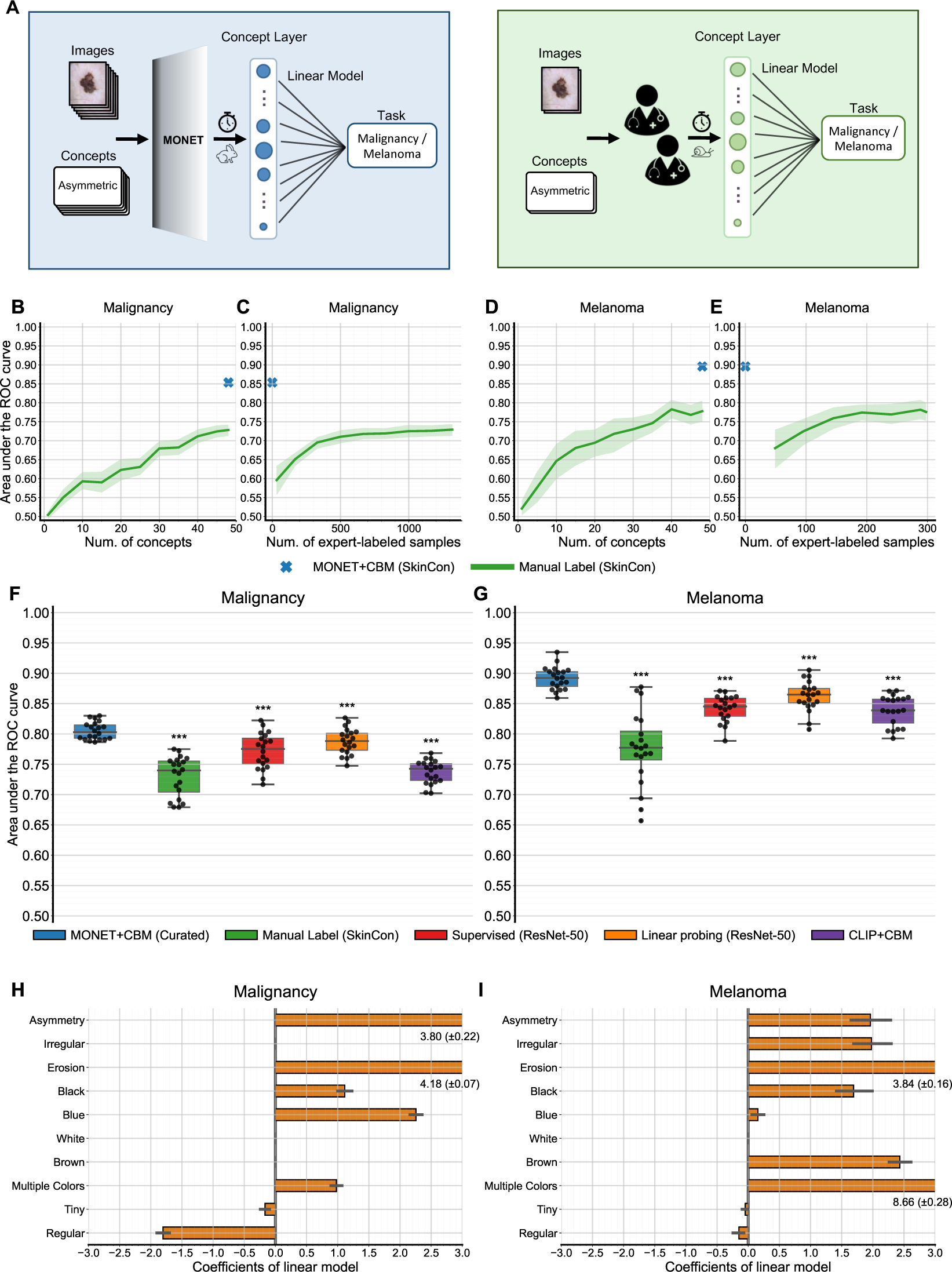
Concept bottleneck model. **(A)** Concept bottleneck model built using concepts generated by MONET (blue). The model first generates concepts using MONET and then predicts disease labels by combining them via a linear model. Concept bottleneck model built using concepts manually labeled by experts (green). The model uses manually annotated concept labels to predict disease labels using a linear model. Manual annotations take a lot longer than concept generation using MONET. **(B)**-**(C)** Performance of a malignancy prediction model trained using manual labels with respect to the number of concepts and the number of expert-labeled samples. **(D)**-**(E)** Performance of a melanoma prediction model trained using manual labels with respect to the number of concepts and the number of expert-labeled samples. **(B)**-**(E)** MONET+CBM is shown as a cross mark because it can utilize all concepts without expert annotation. The shaded area represents the 95% confidence interval. **(F)** Performance comparison of malignancy prediction models. **(G)** Performance comparison of melanoma prediction models. **(F)**-**(G)** Unlike (B)-(E), MONET+CBM uses task-relevant concepts curated by dermatologists. Each dot represents the AUC measure for individual runs with a different train-test split. The box represents the interquartile range with its lower and upper bounds corresponding to the first quartile and third quartile, respectively. *p* values derived from one-sided paired *t*-tests comparing MONET+CBM and other methods are indicated: \**<*0.05, \*\**<*0.01, \*\*\**<*0.001; *n* =20 runs of each method. **(H)** Coefficient of the linear model in MONET+CBM for malignancy prediction. **(I)** Coefficient of the linear model in MONET+CBM for melanoma prediction. **(H)**-**(I)** The error bars present the 95% confidence interval.

We observe that access to a large number of concepts and samples offers performance advantages. Fig. 5B-E compares the performance of MONET+CBM to that of using manual concept annotations. For a fair comparison, we use both methods on the same set of concepts, specifically the 48 concepts in SkinCon. We chose SkinCon concepts because they already have manual annotations provided by experts. For MONET+CBM, we use all training samples and concepts because it can automatically generate concepts without expert annotation. As we increase the number of manually labeled samples used or the number of concepts used, the performance of the CBM created from manual labeling improves. However, even when all manually labeled concepts and training samples are used, the manual approach is not able to match the performance of MONET+CBM, which has access to more samples due to the ability to produce automatic concept labels. As concepts in SkinCon are not annotated for all images, the manual label approach is limited to 1,316 malignancy and 294 melanoma images that have manual concept labels; in contrast, MONET makes use of all 3,968 malignancy and 620 melanoma images in our training set.

We compare the performance MONET+CBM to the other baselines, such as supervised models and CLIP-based CBM, for the same prediction targets (as described in Methods) (Fig. 5F and G). Dermatologists selected 11 target-relevant curated concepts for the bottleneck layer to compare MONET+CBM and CLIP+CBM, which can both flexibly label concepts. Compared to using all 48 SkinCon concepts, the mean AUROC across runs using the 11 curated concepts decreased from 0.854 to 0.805 for malignancy prediction and decreased from 0.896 to 0.892 for melanoma prediction. Still, for both predicting malignancy and melanoma, MONET+CBM outperforms all other baseline methods in terms of the mean AUROC (for malignancy, 0.805 with a standard deviation of 0.014; for melanoma, 0.892 with a standard deviation of 0.019). Out of 20 runs with different random splits of the train and test data, MONET+CBM outperformed all other methods in 15 runs for malignancy prediction and 18 for melanoma prediction, with the linear probing method outperforming in the remaining runs. We also conduct one-sided paired t-tests, comparing the AUROC values of MONET+CBM to those of other methods, where the alternative hypothesis is that MONET+CBM’s AUROC is higher than the other method. In all cases, the resulting p-values are less than 0.001. Thus, MONET’s ability to automatically generate concepts enables the creation of models that are both interpretable and high-performing.

While the concepts used for the concept bottleneck model were selected by dermatologists based on factors that can help predict melanoma, we wanted to check that the way these concepts were used by this model align with established clinical rules for the same task. Fig. 5H and I show the weights of the trained linear classifier corresponding to the concepts used in the bottleneck layer. For the Melanoma target, the results are consistent with the ABCDEs of melanoma [46], which define easily recognizable features—namely, **a**symmetry, **b**order irregularity, **c**olor variation, **d**iameter, and **e**volution—that differentiate malignant melanomas from benign melanocytic nevi. From the concept weights obtained, all concepts that coincide with the ABCDEs get a positive weight as expected, indicating a positive correlation with the melanoma prediction target. The concept “blue” also has a positive weight referring to the dermoscopy concept of blue-white veils observed in melanomas. The concepts “white” and “tiny” get an almost zero weight, while the concept “regular” gets a negative weight, consistent with prior knowledge shared by dermatologists, that regular borders and color indicate a benign lesion. For the malignancy target, no well-defined guidelines exist for deriving concepts; thus, the same concept list as the Melanoma target is used. The results are similar, with a majority of the concepts retaining their directionality, except for increased sparsity in concept weights.

## Discussion

Even with the regulatory approval of AI-supported medical devices, much of the AI pipeline is not transparent - from large-scale datasets that may contain biases to so-called “black-box” models that are not easily audited or interpretable [27]. One approach to improving transparency and trustworthiness is identifying semantically meaningful, human-understandable concepts that are present in datasets or used by models. However, to date, all datasets and methods developed using concepts have relied on human labeling and domain experts, which is not tractable for large-scale, real-world deployment.

Here, we demonstrate the ability to develop automated concept labeling in a medical domain that would usually require domain expertise, and we showcase how these automated concepts can be used to perform tasks for trustworthy AI development at all stages, from developing new models to auditing existing datasets and models. We focus onthe field of dermatology due to the heterogeneity of the image data, the large number of potential concepts, and the ability to validate our methods on existing datasets.

Prior work using image-text models in medicine focused on self-supervised training of diagnostic models that can identify a handful of disease labels, such as in radiology or pathology [29, 47]. However, our challenge is to develop a model that can label a vast number of human-understandable concepts across two image modalities within dermatology: clinical images and dermoscopic images. To solve this challenge, we collect a large number of dermatology image-text pairs from PubMed articles and medical textbooks and train an image-text model, MONET. This dermatology image-test model facilitates automatic generation of concepts, and we show how it can be used to improve the transparency of dermatology AI models. To our knowledge, we are the first to use a large biomedical image-text model to improve the transparency and explainability of medical AI systems.

For a concept generation task where we had domain expert labels as the ground truth, we find that MONET, which requires no domain expert labeling, outperforms the baseline CLIP model and a supervised model trained from domain-expert labeled images. These findings are significant since the bottlenecks of data labeling and domain expertise time can be overcome with image-text models developed from existing medical corpora.

After demonstrating the ability to generate concepts on par with supervised models, we showcase MONET’s ability to facilitate AI auditing and transparency in the dermatology domain. For example, artifacts such as pen markings, stickers, and hair are known to affect dermatology model performance [34, 48]. However, most studies do not assess the influence of artifacts on their models because their datasets are not labeled for these anomalies. We demonstrate MONET’s ability to automatically identify these artifacts, which can be useful for data and model auditing. As an example of how this kind of auditing is useful, we analyzed data from the ISIC 2018 challenge and find that a “red” hue appears more often in benign images for the Medical University of Vienna while images from the Hospital Cĺınic de Barcelona more often have a “red” hue associated with malignant images. This leads to confounding ifa model is trained on one site’s dataset and tested on the other, as we see when we implement MA-MONET for model auditing. These insights derived from using MA-MONET might not be readily achievable via conventional saliency map techniques [43]. For instance, the “red” hue noticed using MA-MONET is not a localized attribute, so a saliency map approach would not necessarily highlight this aspect [43]. Utilizing the insights gained through MONET and MA-MONET, AI model developers can refine data collection and processing and also improve optimization techniques, consequently fostering the development of more reliable and trustworthy medical AI models.

In medicine, inherently interpretable models are of particular interest since they allow physicians to easily decipher the factors influencing a model’s decision. Concept bottleneck models (CBMs) are one such inherently interpretable model but have been limited because they require a priori concept labels, which only a handful of medical datasets contain. MONET overcomes this issue with automatic concept labeling, allowing the creation of CBMs that were not previously possible.

MONET demonstrates the ability to automatically label numerous concepts across heterogeneous disease states and across two modalities (clinical and dermoscopic) in dermatology. A limitation of our experiments is the availability of diverse skin tones in dermoscopic images since no public datasets exist with diverse dermoscopic images [49]. Thus, when assessing MONET on clinical images, we utilize two datasets known to include a diversity of skin tones, Fitzpatrick 17k and DDI, and find that MONET performs well with these datasets.

While MONET covers heterogeneous dermatology data across two modalities, clinical and dermoscopic, future iterations can extend to other forms of medical imaging to improve AI transparency for those use cases. MONET demonstrates that AI transparency and trustworthiness at scale is feasible in a way that was previously impossible: through image-text models tailored to the medical domain of interest.

## Methods

### Dataset

#### Overview

We trained MONET on 105,550 pairs of image and text collected from PubMed articles and medical textbooks [50, 51]. We evaluated MONET using images from the International Skin Imaging Collaboration (ISIC) [21–26], Fitzpatrick17k [52], and Diverse Dermatology Images (DDI) datasets [30].

#### PubMed

The PMC Open Access Subset is a dataset with millions of scientific articles released by PubMed Central (PMC) [50]. First, to find dermatology articles in the dataset, we queried papers in PMC using dermatology-related terms (*i.e.,* dermatology, melanoma, skin cancer), 114 disease labels in the Fizpatrick17k dataset [52], and 48 concept labels in SkinCon [1]. We downloaded 496,510 articles found via this query. In total, the articles contained 3,172,490 figures. Next, we filtered out non-dermatology-related figures (e.g., graphs, illustrations, diagrams, slide images, and X-ray images). To this end, we repeated the process of running a clustering algorithm on the images and manually excluding groups of non-dermatology ones. Specifically, we carried out the following procedure. The clustering features were 50 principal components of the embedding of the penultimate layer of the EfficientNetV2-S model pre-trained on ImageNet [53]. Using the features, we ran a K-means clustering algorithm with the K (*i.e.,* the number of clusters) of 20. For fine-grained filtering, we further applied the K-means algorithm with the K of 20 on each cluster, resulting in400 clusters in each step. For the K-means algorithm, we used the implementation in scikit-learn Python package (ver. 1.2.2) [54]. We manually inspected 50 samples of each cluster and filtered out clusters with non-dermatology images. After going through this step three times, we determined that the remaining clusters contained mostly dermatology images. Post-filtering, 50,265 images remained. Finally, we paired the figure captions to their corresponding images based on the provided XML-formatted file. This file stores the article’s structure with components such as abstract, sections, figures, and figure legends tagged.

#### Textbook

We first extracted images from 55 medical textbooks, yielding a total of 104,223 images. After undergoing the same filtering procedure as we did for PubMed images, 55,285 images remained. The PDF format of the textbooks made matching images with associated text difficult, since PDFs lack the structure information provided by XML formats. To address this issue, we implemented the following procedure. We used PyMuPDF (ver. 1.21.1), an open-source PDF rendering software, to parse the PDF files, extracting text and image objects along with their respective coordinates. Then, we assigned text to images appropriately based on the following criteria. First, we included text that starts with words indicative of figure legends, such as “Fig” or “Figure”. Next, we excluded text based on font and font size. Also, since each textbook maintained a consistent layout for placing figure legends (*i.e.,* legends positioned above or below the figure), we incorporated this into our filtering process. Lastly, from the remaining text, we selected the one closest to the image. We customized the specific parameters (*i.e.,* figure identifier, font, font size, and caption position relative to the image) for each textbook to ensure accurate text-image associations.

#### ISIC

The International Skin Imaging Collaboration (ISIC) archive is a repository of digital skin images, primarily consisting of dermoscopic images, sourced from various institutions. ISIC represents the largest and the most commonly used dataset for the development of dermatology AI [27]. We downloaded 71,671 images in total from all of the ISIC collections, including ISIC Challenge datasets 2016, 2017, 2018, 2019, and 2020 [21–26]. The images have diagnostic attributes, including binary malignancy versus benign labels and 27 granular disease labels. For per-institution analysis, we grouped images by data sources based on the attribution column in the metadata. We selected the two largest cohorts: the Department of Dermatology at the Medical University of Vienna (9,873 samples) and the Department of Dermatology at the Hospital Cĺınic de Barcelona (12,302 samples).

#### Fitzpatrick17k

Since the PubMed and textbook datasets contain clinical (*i.e.*, non-dermoscopic) images, for evaluation purposes, we required additional clinical images with ground-truth annotations. As the first of these datasets, we chose Fitz-patrick17k[52], which contains dermatological images collected from online dermatology atlases accompanied by disease annotations and Fitzpatrick skin type labels. To reduce the impact of artifacts in the images, we filtered the dataset to exclude images with visible patient clothing, visible anatomy (*e.g.*, fingers, ears, eyes, etc.), or other elements except for lesions and background skin. To filter these images, we first manually annotated 10% of the full dataset (1,657 of 16,577 images), marking each image as include or exclude, then trained a machine-learning model to classify the remaining 90% of images. In particular, we fine-tuned a DenseNet-121[55] (pre-trained on ImageNet) to predict exclusions using our 80% of our hand-labeled data, then chose an operating point to maximize the F1 score (maximum = 0.81) on the remaining 20% of the hand-labeled data. This filtering resulted in a total of 4,951 images, incorporating both those that passed the classifier’s screening and 462 images from our hand-labeled set. Fitzpatrick17k contains near duplicate images with slight differences in angle or cropping; we filter for duplicate images to prevent overlap between the train and test sets for the concept bottleneck model experiments. To measure the distance between images, we obtained the 50 principal components of the embedding of the penultimate layer of the EfficientNetV2-S model (pre-trained on ImageNet) [53]. Then, we calculated cosine similarity between the 50 principal components of EfficientNet embedding. To rigorously filter out duplicates, we used a loose threshold (cosine similarity = 0.9) and manually identified any false positives. In total, among the 4,951 images in Fitzpatrick17k “clean” set, we identified 523 sets of duplicate images, with some sets containing up to 6 duplicates. When selecting which images to keep among the duplicates, we prioritized keeping those images with SkinCon annotations. After this filtering, we had a total of 4,386 images. Lastly, we excluded 62 images that were marked as ‘Do not consider this image’ (*i.e.,* images of low quality or considered not appropriate) in the SkinCon dataset. This led to a final dataset containing 4,324 images.

Additionally, for melanoma prediction tasks, amongst the 113 fine-grained diagnosis labels, we further refined the data to include only melanomas and melanoma look-alikes, such that the data mirrors a well-defined clinical task. In line with the disease filtering criteria outlined by Degrave et al. [43], we included melanomas, benign melanocytic lesions, seborrheic keratoses, and dermatofibromas, resulting in a total of 500 images.

#### DDI

As a second set of clinical images with ground truth labels, we chose the Diverse Dermatology Images (DDI) dataset. DDI contains 656 clinical images of diverse skin tones, obtained from Stanford Clinics [30], accompanied by annotations of Fitzpatrick skin type and histopathologically proven diagnoses. Again, we excluded 20 images that were marked as ‘Do not consider this image’ in the SkinCon dataset, resulting in the final dataset of 636 images. For melanoma prediction tasks, we narrowed the dataset to include only melanomas and melanoma look-alikes, resulting in a total of 275 images, in accordance with the approach taken by Degrave et al. [43]. Among the 78 fine-grained diagnosis labels in DDI, the melanoma category comprises the general label “melanoma” as well as the more specific labels acral-lentiginous melanoma, melanoma *in situ*, and nodular melanoma. Melanoma look-alikes consist of acral melanotic macule, atypical spindle cell nevus of reed, benign keratosis, blue nevus, dermatofibroma, dysplastic nevus, epidermal nevus, hyperpigmentation, keloid, inverted follicular keratosis, melanocytic nevi, nevus lipomatosus superficialis, pigmented spindle cell nevus of reed, seborrheic keratosis, irritated seborrheic keratosis, and solar lentigo.

#### SkinCon

SkinCon is at present the most comprehensive dataset on dermatology concepts [1]. The dataset features 48 concepts, curated by board-certified dermatologists, that are frequently used to describe skin lesion attributes such as shape, size, color, and texture. The dermatologists manually annotated the ground-truth labels for these concepts on 3,230 images from the Fitzpatrick17k dataset, which originally consisted of 16,577 images, and all 656 images from the DDI dataset. Of the 4,324 images in the Fitzpatrick17k dataset we obtained after filtering, 1,009 had SkinCon annotations. Among the 500 images in the dataset used for the melanoma prediction task, 95 had annotations.

#### MONET

Formally, let *f*_image_ : *X*_image_ *→* R*^d^* be the MONET image encoder and *f*_text_ : *X*_text_ *→* R*^d^* be the MONET text encoder. Given a dataset of paired images *I ∈ X*_image_ and text descriptions 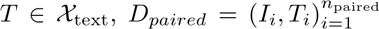, our goal is to train the two encoders such that the distances between pairs of embeddings *dist*(*f*_image_(*I_i_*), *f*_text_(*T_j_*)) reflect the semantic similarity between *I_i_* and *T_j_* for all *i, j ≤ n*_paired_.

#### Architecture

We use the vision transformer architecture, ViT-L/14, as our image encoder [56]. This encoder takes an input image of size 224 x 224 and outputs a 768-dimensional embedding. For the text encoder, we use a transformer architecture with 12 self-attention layers. It takes tokenized text with a maximum limit of 77 tokens as input and outputs a 768-dimensional embedding. We use the same architecture as CLIP [20] to take advantage of the weights from pre-trained models.

#### Preprocessing

To meet the input requirements for the encoder architectures, we process image and text inputs as follows. Each input image is re-sized and center-cropped to be 224×224 dimensions. It is then normalized using the mean and standard deviation used in CLIP [20]. Throughout the training phase, we applied standard data augmentation steps instead, such as random resized crops, vertical flips, horizontal flips, and color jittering for brightness, contrast, and saturation. For each input text, we apply tokenization using lower-cased byte pair encoding [57]. In cases the text encountered during training was longer than the text encoder’s maximum token limit of 77, we split the text into sentences and chose half of them from the beginning. We repeated this process until the token count was reduced to fewer than 77.

#### Training

We use cosine similarity as the distance metric. Both encoders are jointly trained to maximize the cosine similarity between the image and text embeddings of the correct pairs while minimizing the cosine similarity between the embeddings of incorrect pairings. To this end, we use a symmetric cross-entropy loss on cosine similarity scores; after calculating the cosine similarities between embeddings, we scale them by a temperature parameter *λ* and normalized them into a probability distribution with the softmax function. The temperature parameter *λ* was also updated during training. We optimize the loss using the Adam optimizer [58] with a cosine learning rate schedule for 10 epochs. This implementation detail follows that of CLIP [20].

For hyper-parameter tuning, we split the dataset into training and validation sets and find hyper-parameters that result in the best validation loss; we use validation loss for hyper-parameter tuning because there is no large-scale ground truth label for evaluating concept generation performance. We tune the hyper-parameter of batch size (128, 256, 512, 1024) and learning rate (1e-3, 1e-4, 1e-5, 1e-6). We find that the larger batch size results in lower validation loss until a batch size of 512 is reached. We also find that the learning rate of 1e-5 leads to the lowest validation loss. Using the tuned hyper-parameters, we train the model on the whole dataset for 10 epochs. We use 6 Nvidia A40 GPUs with data parallelism. Model training takes 1 hour and 40 minutes.

### Automatic concept generation

During the training procedure of image and text encoders, an image and a text from the same pair are forced to be close to each other in the embedding space, while ones from different pairs are forced to be far apart. After training, MONET can measure the proximity between an image and any arbitrary text. We use this capability to automatically generate concepts for images.

To generate a concept *c* for a given batch of N images *I*_1_, *I*_2_, · · ·, *I_N_*, we first compute the image embeddings *f*_image_(*I*_1_), *f*_image_(*I*_2_), · · ·, *f*_image_(*I_N_*) using the image encoder *f*_image_. We also compute the concept prompt embedding *f*_text_(*T_c_*) and reference prompt embedding *f*_text_(*T_r_*) using the text encoder, where *T_c_* is a concept prompt (*e.g.,* “This is a skin image of”) and *T_r_* is a reference prompt (*e.g.,* “This is a skin image of”). Supplementary Table 4 shows the terms used for each concept for filling templates. Next, we calculate the cosine similarity between image embeddings and prompt embeddings. When multiple terms are used for each concept, we calculate the cosine similarity for each term and average them. Finally, we obtain concept presence score *p_i,c_* that represents the degree to which a concept is present in the image as follows:

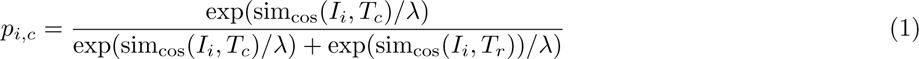

where sim_cos_(*·*) is the cosine similarity score between image embeddings and text embeddings, sim_cos_(*I_i_, T_c_*) = 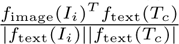, and *λ* is the temperature parameter learned during the training. We normalize by reference prompt to remove the effect of templates being used. Further, we use multiple templates to minimalize the effects of templates. For clinical images, we used templates: “This is skin image of *{}*”, “This is dermatology image of *{}*”, and “This is image of *{}*”. For dermoscopic images, we used the templates “This is dermatoscopy of *{}*” and “This is dermoscopy of *{}*”. We use concept presence scores averaged across different templates in the end.

#### Quantitative evaluation

We use 1,645 images with SkinCon labels from Fitzpatrick17k and DDI datasets for the task of predicting SkinCon concepts. We use 4,324 images from Fitzpatrick17k and 636 images from DDI datasets for the task of predicting disease labels, respectively. We compare the performance of MONET’s concept generation to that of a supervised learning approach, training a ResNet-50 model [28], and to that of a pre-existing image-text model CLIP [20]. MONET and CLIP do not require additional training to perform these tasks; the output from MONET and CLIP models is obtained via the automatic concept generation procedure described above. In contrast, we need to train a supervised learning model. We train the model using a standard training recipe as follows. We initialize the model using ImageNet pretrained weights. We then replace the last layer of the model with a new MLP layer that matches the dimension of the prediction target; for SkinCon concepts, we train each concept one by one (the dimension is 1), and for disease labels, we train disease labels considered all at once (the dimension is 113 for Fitzpatrick17k and 78 for DDI). We finally train the model using cross-entropy loss for 20 epochs. We use the Adam optimizer [58] with a ReduceLROnPlateau learning rate scheduler implemented in Pytorch (ver. 1.13.0); the initial learning rate is 1e-3, and it is reduced based on validation loss with the patience parameter of 2. Also, we use EarlyStopper implemented in PyTorch, which stops the training when the validation loss does not improve 5 times. The available data for each task is split into train/test sets with a ratio of 4:1, and 20% of the train set is left for calculating validation loss. While for MONET and CLIP, we calculate AUROC across all available samples in one go, for the ResNet-50 model, we repeat the evaluation with 20 different train-test splits and calculate the average AUROC for each target to leverage all samples fully.

### Data auditing

#### Concept differential analysis

MONET’s ability to map images and texts onto the co-embedding space enables us to describe the different characteristics between two sets of images in natural language. Assume we have two sets of images, denoted as **I**_+_ = *I*_1_, *I*_2_, …, *I_N_* + and **I***_−_* = *I*_1_, *I*_2_, …, *I_N_−*, and a list of concepts we want to investigate [*c*_1_, *c*_2_, …, *c_Nc_*]. We first obtain the prototype embedding of each image set by computing an average of normalized image embeddings, 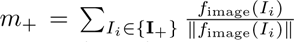 and 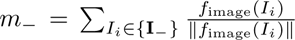. Then, we calculate the displacement vector from *m*_-_ to *m*_+_ by subtracting out the two prototype embedding *m*_Δ_ = *m*_+_ *-m_−_*. Finally, we get a differential concept expression score by computing the dot product between the prototype and normalized embeddings of concept prompt 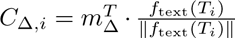. This score measures how much more each concept is differentially expressed in *S*_+_ than in *S_−_*. A similar technique for converting a set of images to text has been previously used by Eyuboglu et al. [13].

#### Benchmark analysis

To perform a benchmark study on concept differential analysis, we construct synthetic data using ground-truth concept labels in the SkinCon dataset. For each concept in SkinCon, we create a dataset split into two groups: one with 100 images, many of which are associated with the concept, and another with 100 images, many of which are not associated with the concept. We use the noise level parameter to control the degree to which the concept is correlated with the grouping; it indicates the probability that images are randomly sampled from the opposite group. We run simulations 20 times for each combination of parameters with different random seeds.

### Model auditing

#### Model auditing with MONET

We can use MONET to automatically detect semantically meaningful medical concepts that lead to model errors. Model auditing with MONET (MA-MONET) starts by sorting images from a test set into groups based on their visual similarity. To this end, we run the K-means clustering algorithm implemented in the scikit-learn Python package (ver. 1.2.2) [54, 59]. We use 50 principal components of the embedding of the penultimate layer of the EfficientNetV2-S model (pre-trained on ImageNet) [53] as clustering features. Next, we calculate the accuracy across all samples and also per cluster; for thresholding the probability output from the trained classifier, we choose an operating point that maximizes the F1 score. Following this, we identify medical concepts for the “low-performing” cluster; we define low-performing clusters as ones with accuracy lower than overall accuracy. Each low-performing cluster is compared to a high-performing counterpart containing similar images to understand what differentiates them; among the clusters that perform better than the overall accuracy, we choose one whose centroid is closest in Euclidean distance to the low-performing cluster. We conduct a concept differential analysis between the high and low-performing clusters to pinpoint concepts that are more presented in the low-performing cluster. If two visually similar clusters (one high-performing, the other low-performing) differ in terms of a few concepts, these differing concept terms can be hypothesized as leading to high error rates. We then filter out concepts with a concept presence score below 0.5 in the low-performing. Finally, we obtain a ranked list of medical concepts identified by MONET that differentiate the two clusters.

#### Benckmark analysis

For benchmarking analysis, we use a situation where the ground truth (*i.e.,* the concepts leading to model error) is already known. We create a training dataset with spurious correlations from the Fitzpatrick17k and DDI datasets: 500 malignant images that feature a particular SkinCon concept, while the 500 benign images do not. For the test set, we reverse the correlation; 500 sampled benign images have the SkinCon concept, while 500 sampled malignant images do not. For concepts of spurious correlation, we use 5 concepts that remain after filtering out concepts with fewer than 30 samples in each category required for creating the confounded training and test sets (*i.e.,* malignant–with concept, malignant–without concept, benign–with concept, and benign–without concept): “crust”, “hyperpigmentation”, “plaque”, “erythema”, and “papule”. For each of the 5 selected concepts, we repeat this analysis 20 times with different random seeds varying the training and test sets. Consequently, we conduct analysis for a total of 100 settings. In addition to the concept we intentionally introduce as a confounder, there are other concepts that also inadvertently become confounders. For example, when we had “papule” to be associated with malignancy in the train set, “plaque” was associated with benign images in the training set. In such cases, we define all of them as the ground truth. On average, there are two concepts across all 100 test settings. We consider the spurious correlations are recovered if the top-N concepts identified by MA-MONET include at least one of these ground truth concepts across all low-performing clusters.

### Building inherently interpretable neural network

#### Concept Bottleneck Model

Concept Bottleneck Models (CBMs) [15] are inherently interpretable models that identify the importance of each concept for the classifier’s prediction. They use a bottleneck layer to extract compact and discriminative representations of the input data. The bottleneck layer, typically composed of a small number of units, imposes a constraint on the amount of information transmittable through the network, forcing it to make predictions by using interpretable features that align well with the users’ expectations. This technique can be used to reduce the dimensionality of the data and improve the efficiency of the model while preserving its predictive power. CBMs have been successfully applied to a wide range of tasks, including image and video classification [60, 61], natural language processing [62], and being applied in different medical settings [1, 63, 64]. However, a caveat is that these models need a large set of concept annotations to perform well, and collecting these labels is laborious and time intensive.

MONET lets us automatically generate concepts for images that can be used to scale to a large concept dataset with an arbitrary number of concepts. Specifically, MONET helps to create the bottleneck layer, denoted by *b_c_* : *X*_image_ *→* R*^Nc^*, that maps an input image *I_i_* to a vector of dimension *N_c_*, the number of concepts, where each dimension corresponds to one of the *N_c_* concepts. An interpretable linear model is then trained on the prediction target to get importance scores for each concept corresponding to the trained model weights.

To create the bottleneck layer, we start with a concept list [*c*_1_, *c*_2_, …, *c_Nc_*], chosen with guidance from our dermatologist collaborators, containing concepts that are predictive of the target. Ideally, the bottleneck layer is binarized using the concept labels. However, we lack access to the concept annotations, and thresholding the similarity score of each concept with the input image is non-trivial. Instead, for each concept *c_j_*, we curate a set of reference concepts [*r_j_*_1_, *r_j_*_2_, …, *r_jNj_*] where *N_j_* is the number of reference concepts for concept *c_j_*. Each reference concept is selected such that it is sufficiently far from the concept of interest in the representation space while being closer to the other reference concepts. We do this by choosing antonyms of the concept of interest as the reference concepts.

Once the set of reference concepts is created, MONET calculates the similarity scores of the input image to the concept of interest and the corresponding reference concepts. The former is then normalized by taking a softmax with the reference concept scores. The resulting normalized score, 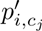, is then used in the bottleneck node for that concept, as shown in Equation 2.

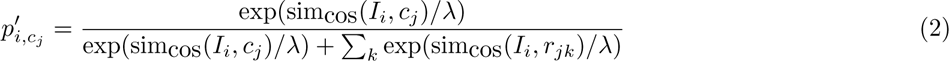

where sim_cos_(·) is the cosine similarity score obtained using MONET, and *λ* is a temperature parameter used to magnify the differences in similarity scores. *λ* is manually tuned to the value that performs the best on the train set. Once the bottleneck layer is created, we train a simple linear classifier on the prediction target using stochastic gradient descent. Specifically, for a classifier *f* and a given sample *x* ∈ R*^Nc^*, the prediction obtained is *w^T^ f* (*x*) + *b*. We apply L1 regularization to favor sparsity in the trained model weights and make the model more interpretable. Once the linear classifier is trained, the learned weights *w* can be analyzed to understand the importance of each concept for the prediction target.

To demonstrate the efficacy of MONET, we use two different prediction targets: (1) Melanoma vs. Melanoma look-alike, and (2) Malignant vs. Benign. We differentiate between these two targets since all melanomas are malignant, but not all malignant lesions are melanoma. For this experiment, we use the clean Fitzpatrick 17k[52] and DDI[30] datasets. We use 80% of the data for training and reserving the rest for testing. To create the bottleneck layer, we use 11 concepts that are known to be correlated to the prediction targets; specifically, we use the ABCDEs of melanoma [46] as a guideline to compile the list of concepts for the bottleneck layer. Supplementary Table 5 lists these concepts along with the reference concepts used for normalization. MONET’s ability to automatically generate concepts for images lets us easily add more data or concepts as needed without any manual annotations.

We compare MONET+CBM to several other baseline methods of obtaining target predictions from input images:

- **Vanilla CLIP+CBM:** We use an out-of-the-box CLIP model to create the bottleneck layer and trains a linear classifier, similar to MONET+CBM. The only difference is that the vanilla CLIP model is not fine-tuned on dermatology images and thus lacks the context of the setting in which we run the experiment; as a result, it cannot adequately capture the semantic differences between technical dermatology terms.
- **Supervised:** We train a deep learning model using the standard fully supervised approach without incorporating concepts. We use ResNet-50 pre-trained on the ImageNet where the last classification head was replaced to match the dimension of the prediction target. We train the model end-to-end to classify the input images into the target classes. The implementation details are the same as described in the Qualitative evaluation subsection under Automatic concept generation. We only change the maximum training epoch from 20 to 50
- **Linear Probing** We use the representation of the penultimate layer of ResNet-50 pre-trained on ImageNet as the input for a linear model. The difference with supervised is that during the training, the backbone of ResNet-50 is frozen.
- **Manual Labeling** We use the SkinCon dataset [1], which applies concept annotations covering 48 concepts for 3230 images from the Fitzpatrick 17k dataset to create the bottleneck layer. These concepts were chosen by two board-certified dermatologists considering the clinical descriptor terms used to describe skin lesions.

## Data Availability

PMC Open Access Subset is publicly available from https://www.ncbi.nlm.nih.gov/pmc/tools/openftlist/. Evaluation datasets are all publicly available and can be accessed from: ISIC (https://challenge.isic-archive.com/data/), Fitzpatrick17k (https://github.com/mattgroh/fitzpatrick17k), and DDI(https://stanfordaimi.azurewebsites.net/datasets/35866158-8196-48d8-87bf-50dca81df965).

## Code availability

The code used in our analysis is available at https://github.com/suinleelab/MONET. It includes various scripts for data collection and preprocessing, training the MONET model, and conducting benchmark studies. Also, it provides the MONET model weights.

## Acknowledgements

We thank Chris Lin and other people in the Lee Lab for helpful discussions.

## Funding

C.K., S.U.G., A.J.D., and S.-I.L. were supported by the National Science Foundation (CAREER DBI-1552309 and DBI-1759487) and the National Institutes of Health (R35 GM 128638 and R01 AG061132). C.K. was supported by the Asan Foundation Biomedical Science Scholarship. R.D. was supported by the National Institutes of Health (5T32 AR007422-38) and the Stanford Catalyst Program.

## Ethics declarations

### Competing interests

R.D. reports fees from L’Oreal, Frazier Healthcare Partners, Pfizer, DWA, and VisualDx for consulting; stock options from MDAcne and Revea for advisory board; and research funding from UCB.

---

**Supplementary Fig. 1.**
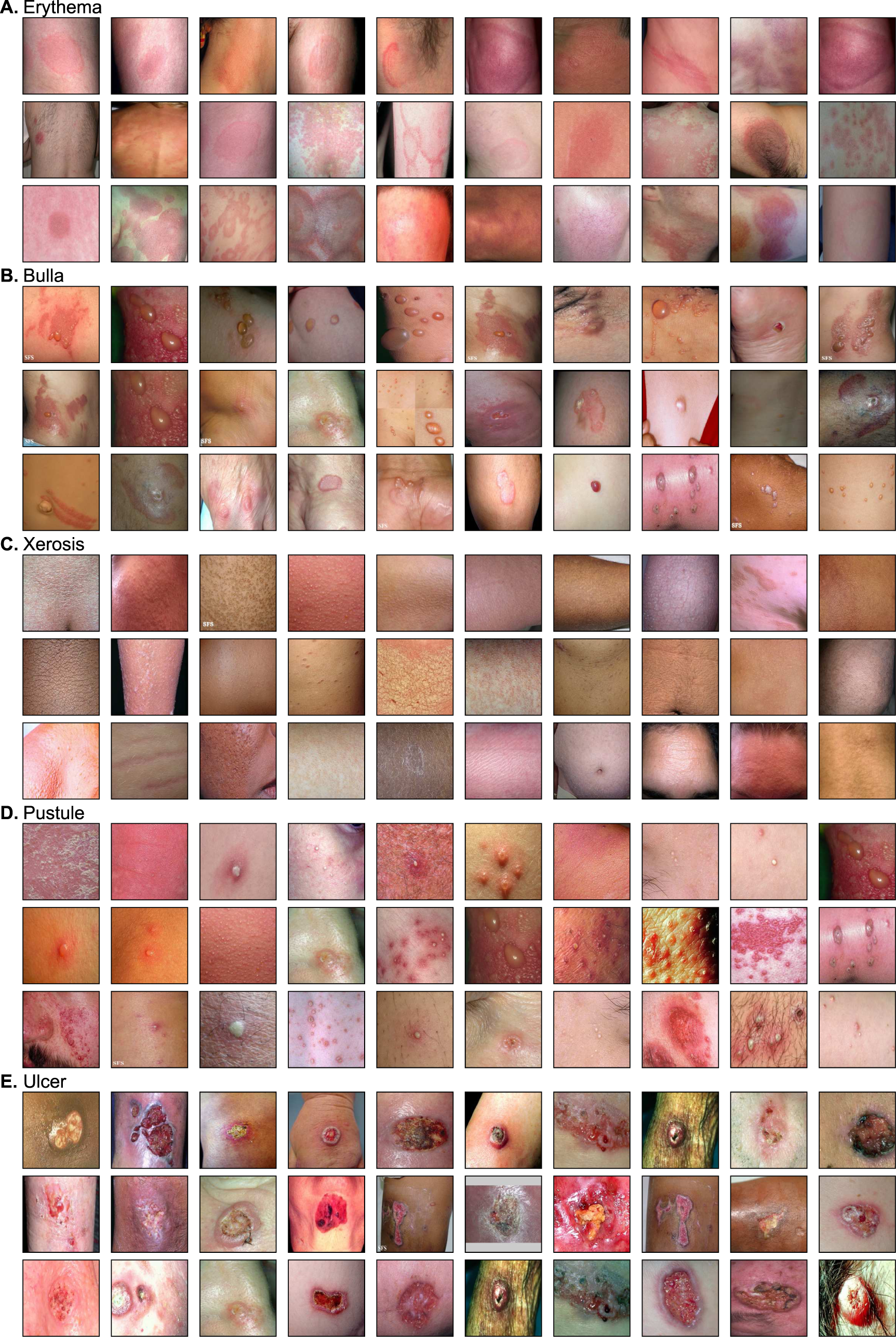
Clinical images with high concept presence scores calculated using MONET. We show the top 30 images for each concept. **(A)** Erythema. **(B)** Bulla. **(C)** Xerosis. **(D)** Pustule. **(E)** Ulcer.

**Supplementary Fig. 2.**
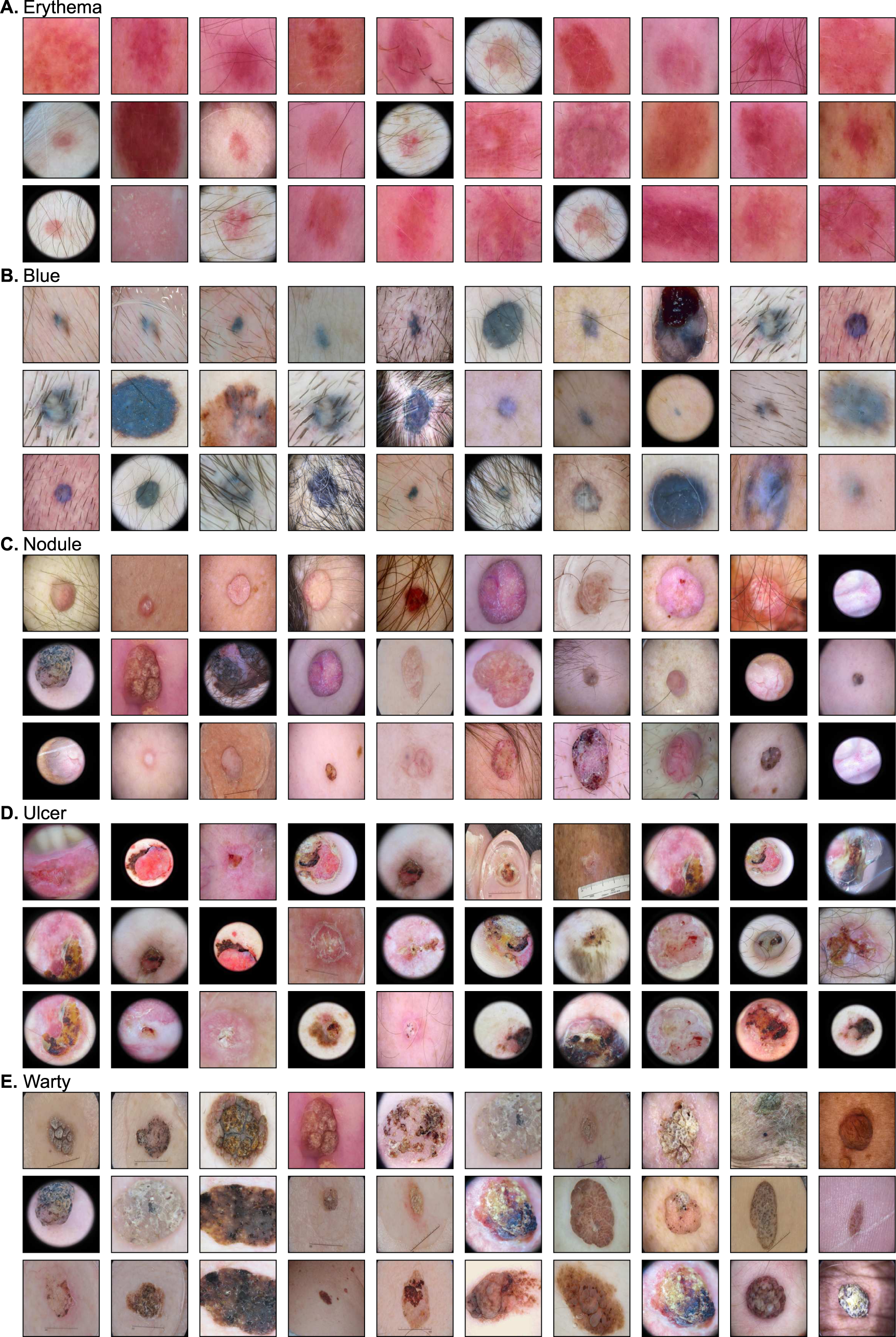
Dermoscopic images with high concept presence scores calculated using MONET. We show the top 30 images for each concept. (A) Erythema. (B) Blue. (C) Nodule. (D) Ulcer. **(E)** Warty.

**Supplementary Fig. 3.**
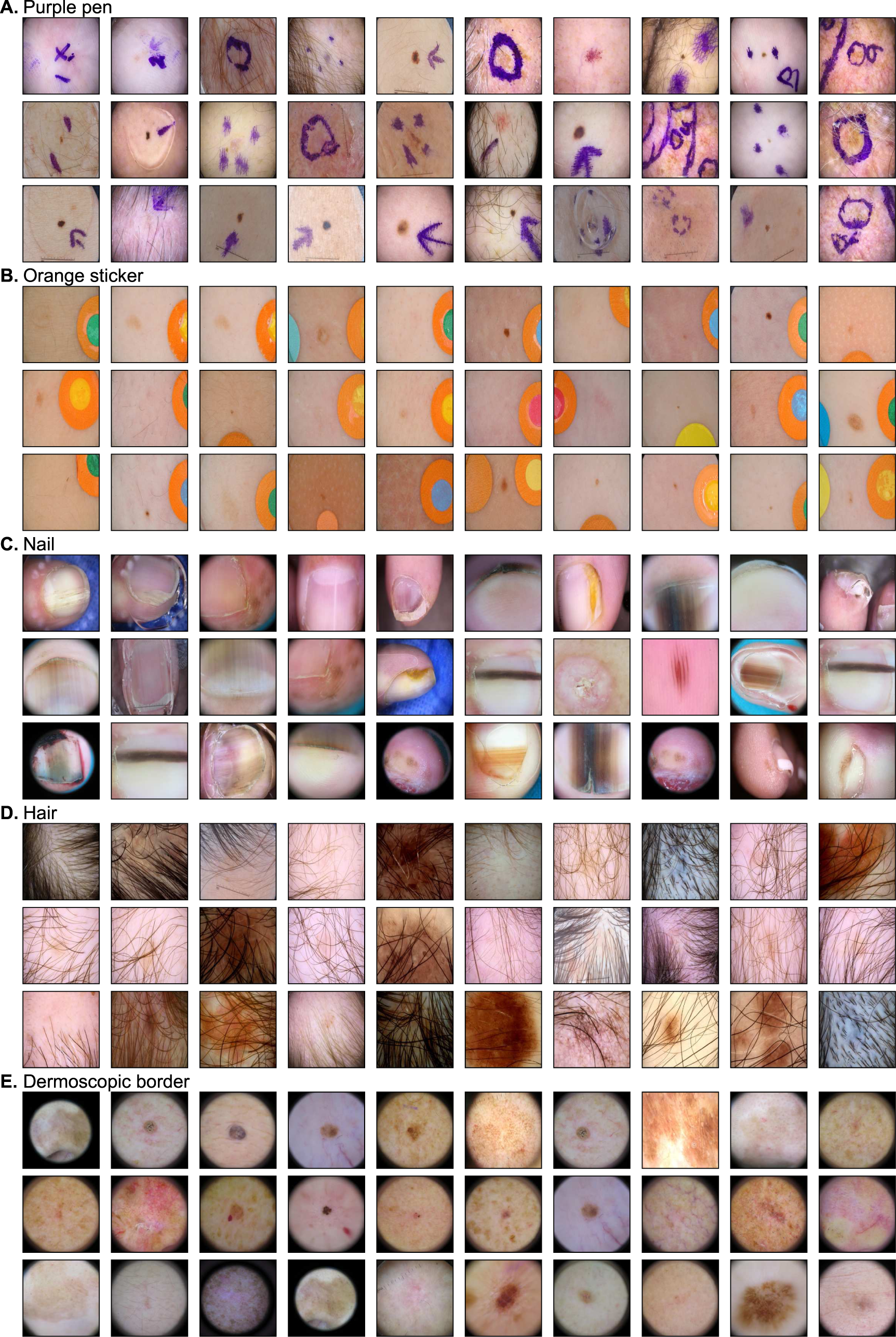
Dermoscopic images with artifacts as determined by high concept presence scores calculated using MONET. We show the top 30 images for each artifact. (A) Purple pen. (B) Orange sticker. (C) Nail. (D) Hair. (E) Dermoscopic border.

**Supplementary Fig. 4.**
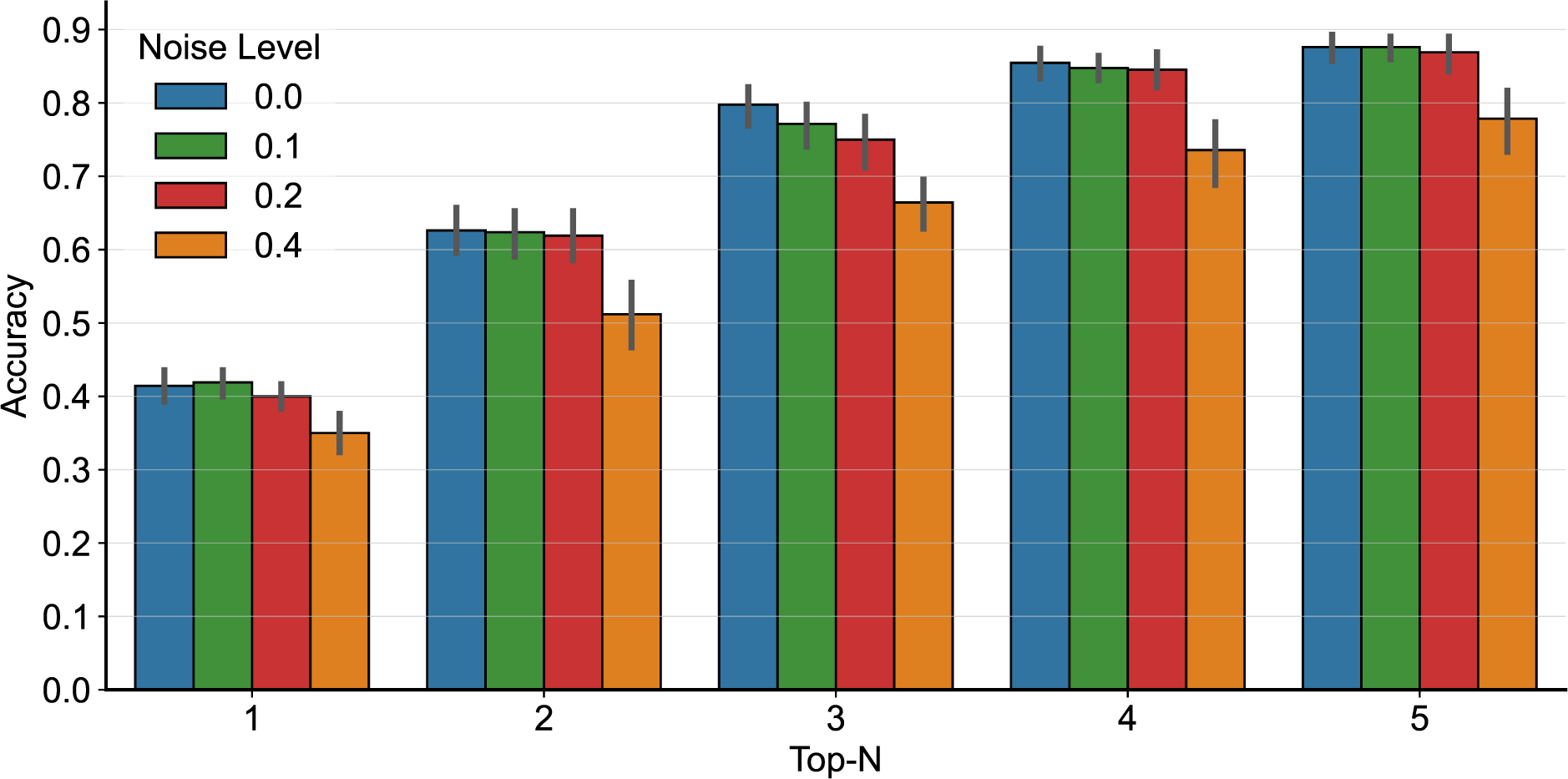
Accuracy of concept differential analysis. We perform a benchmark analysis to assess MONET’s ability to identify presented concepts correctly. To do this, we generate two paired datasets with known ground truth (i.e., a specific concept is differentially presented) and conduct concept differential analysis on these datasets, letting us determine how accurately the analysis recognizes the intended concept. This experiment is conducted on 21 out of 48 concepts from SkinCon that remained after excluding those with fewer than 30 positive examples. For each concept, we sample a set of 100 images where a concept is highly presented and another set of 100 images where a concept is highly absent from Fitzpatrick17k and DDI datasets, with replacement. Additionally, we varied the noise parameters (0, 0.1, 0.2, and 0.4), which control how correlated the concept is to each grouping. For example, noise = 0.1 means that in the “concept present” set, 90% of the images have the concept, while in the “concept absent” set, only 10% of the images have the concept. For each combination of settings (i.e., 21 intended concepts and 4 noise levels), we repeat this evaluation 20 times with different random seeds. The error bars represent the 95% confidence interval.

**Supplementary Fig. 5.**
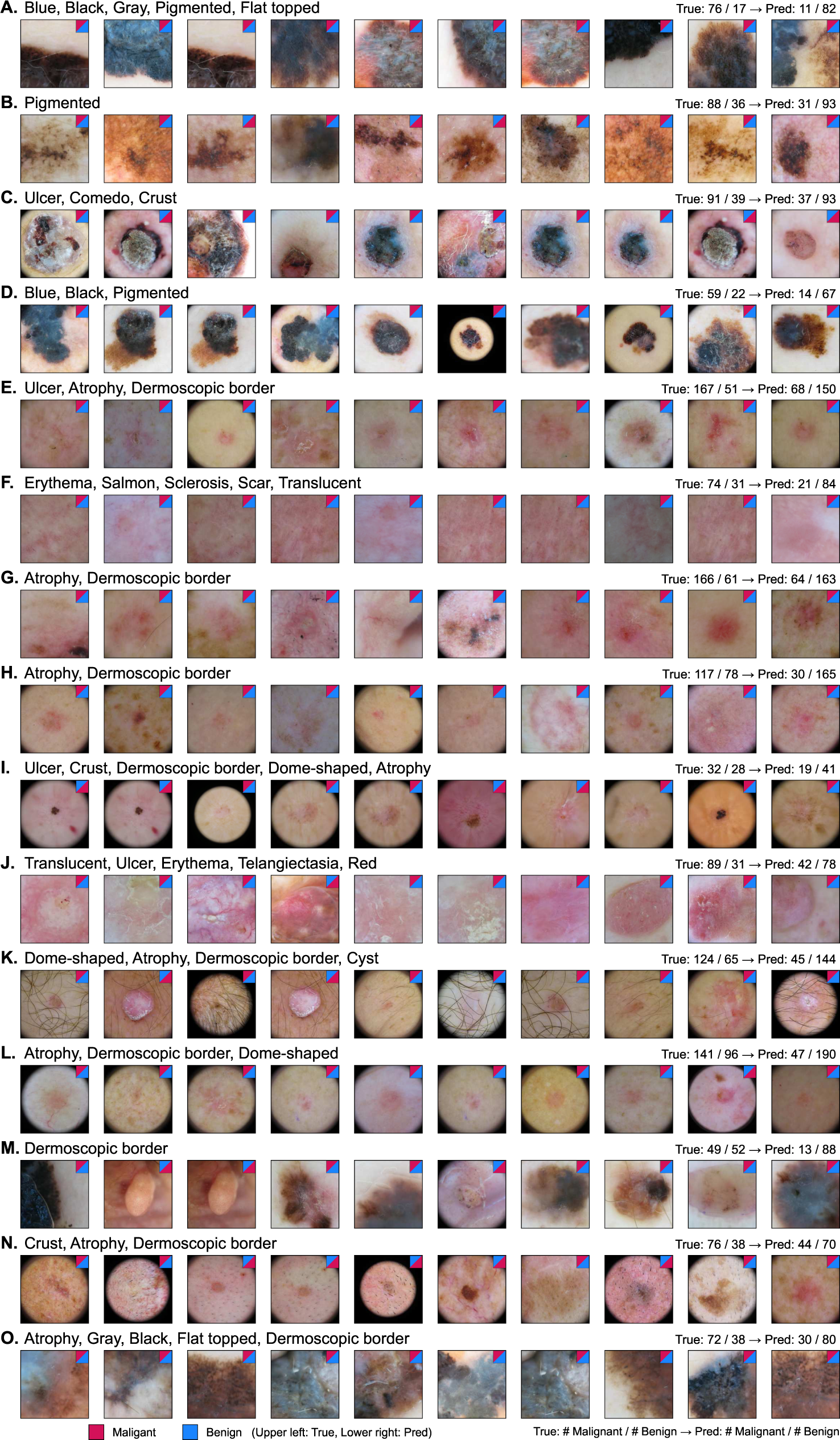
Concept-level model auditing. We train a model on the Med U. of Vienna dataset and test it on the Hosp. Barcelona dataset. Each row displays one of the top 15 clusters, sorted by high error rates. For each cluster, we show the misclassified images and the corresponding concepts associated with errors. We represent the true and predicted labels for each image by the color of the upper left and lower right triangles in the small box, respectively. The numbers at the top right compare the number of malignant and benign samples for the true and predicted labels. The 10 misclassified images shown for each are selected based on the average concept presence of the identified concepts.

**Supplementary Fig. 6.**
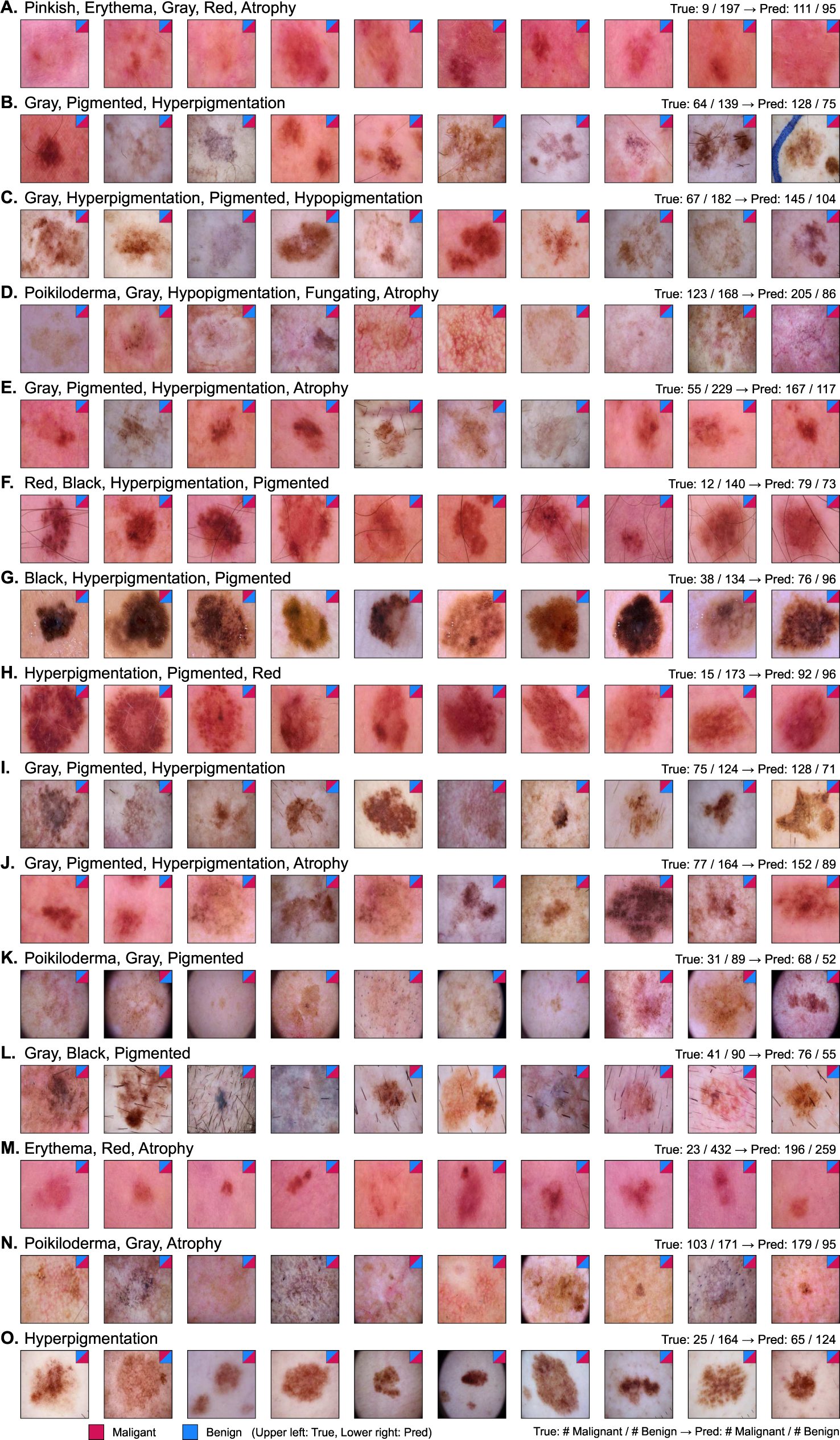
Concept-level model auditing. We train a model on the Hosp. Barcelona dataset and test it on the Med U. of Vienna dataset. Each row displays one of the top 15 clusters, sorted by high error rates. For each cluster, we show the misclassified images and the corresponding concepts associated with errors. The 10misclassified images shown for each are selected based on the average concept presence of the identified concepts.

**Supplementary Table 1.** Images excluded from figures. We exclude 4 images inappropriate for public display due to the inclusion of sensitive body parts, such as genitals, breasts, and buttocks, from Fig. 2 and Supplementary Fig. 1. Their file names, as well as the dataset they belong to, are noted.

| Concept | File name (Dataset) |
| --- | --- |
| Erythema | 58b4bc079ca94e6e9377a42ca7564b40.jpg (Fitzpatrick17k) |
|  | 720cf31558966c82c118ab75b50632eb.jpg (Fitzpatrick17k) |
|  | 5f046cda32a3cc547205662e7be774f9.jpg (Fitzpatrick17k) |
| Ulcer | d8bf377acc45a3beb0c6e81bf7ac1ff5.jpg (Fitzpatrick17k) |

**Supplementary Table 2.** Performance of MONET in annotating disease labels as compared to baselines. We use disease labels in the clinical image datasets, Fitzpatrick17k and DDI datasets, as ground truth. We exclude any with less than 30 positive examples, leaving 59 labels in Fitzpatrick17k and 6 labels in DDI for our analysis. We use 4,324 samples from Fitzpatrick17k and 636 samples from DDI. The baselines are CLIP, an image-text model not specifically trained on dermatology images, and the ResNet-50 model trained on ground truth labels in a fully supervised manner. The numbers in parentheses represent the count of concepts for which the method achieves an AUROC over 0.7 over the total number of diseases examined.

| Method | Mean AUROC |  |
| --- | --- | --- |
|  | Labels in Fitzpatrick17k | Labels in DDI |
| MONET | 0.830 (52/59) | 0.701 (4/6) |
| CLIP | 0.680 (28/59) | 0.595 (0/6) |
| Fully supervised (ResNet-50) | 0.856 (58/59) | 0.700 (2/6) |

**Supplementary Table 3.**
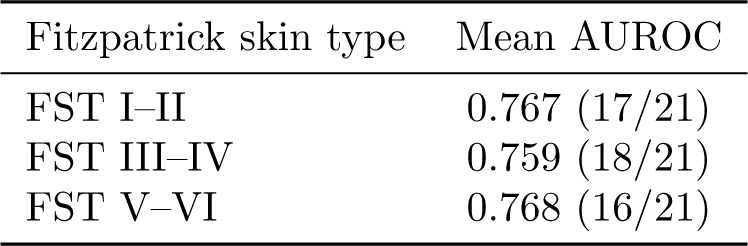
Evaluation of MONET’s concept generation performance per skin tone. We calculate AUROC metrics per each Fitzpatrick skin type (FST) separately: FST I–II (light skin tone, *n* = 717), FST III–IV (intermediate skin tone, *n* = 607), and FST V–VI (dark skin tone, *n* = 283). The numbers in parentheses represent the count of concepts for which the method achieves an AUROC over 0.7 over the total number of concepts examined.

| Fitzpatrick skin type | Mean AUROC |
| --- | --- |
| FST I–II | 0.767 (17/21) |
| FST III–IV | 0.759 (18/21) |
| FST V–VI | 0.768 (16/21) |

**Supplementary Table 4.**
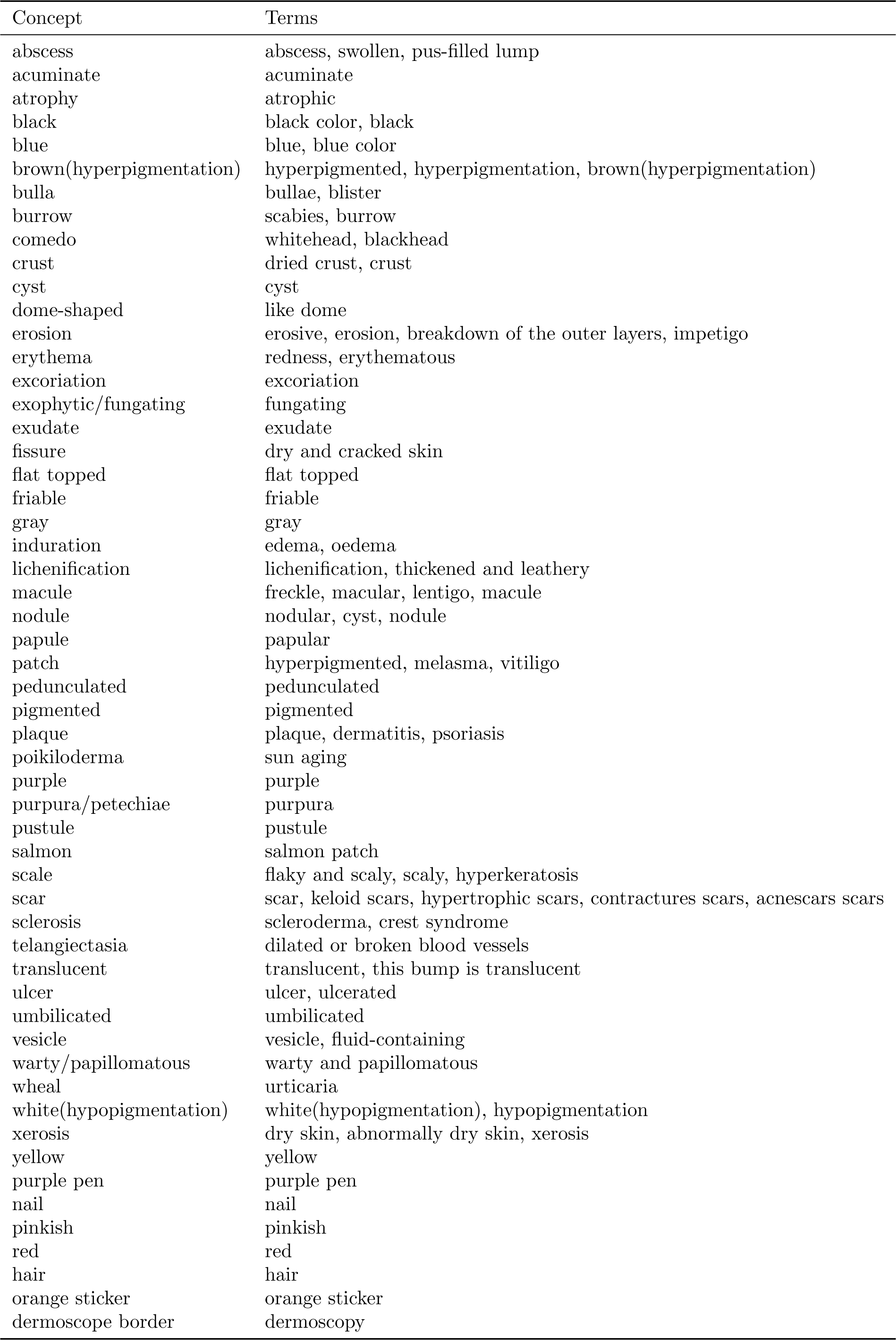
*|* Terms used to generate concept prompts

**Supplementary Table 5.** *|* Concepts used in the bottleneck layer for the Concept Bottleneck Model

| Concept of Interest | Reference Concepts |
| --- | --- |
| Asymmetry | Symmetry, Regular, Uniform |
| Irregular | Regular, Smooth |
| Blue | Green, Red |
| White | Black, Colored, Pigmented |
| Brown | Pale, White |
| Black | White, Creamy, Colorless, Unpigmented |
| Erosion | Deposition, Buildup |
| Multiple Colors | Single Color, Unicolor |
| Tiny | Large, Big |
| Regular | Irregular |

